# Risk of Serious Infections in Patients with Psoriasis on Biologic Therapies: An Updated Systematic Review and Meta-Analysis

**DOI:** 10.1101/2021.08.27.21262722

**Authors:** L. Manounah, Z.Z.N. Yiu, S.K. Mahil, L.S. Exton, M.C. Ezejimofor, A.D. Burden, R. Murphy, C.M. Owen, R. Parslew, R.T. Woolf, C.H. Smith, M.F. Mohd Mustapa

## Abstract

**Background:** The increase in the number of available biologic agents for psoriasis has led to an urgent need to evaluate their associated risk of serious infections to help inform clinical decisions.

**Methodology:** We systematically searched PubMed, Medline, Embase, and Cochrane databases for biologics targeting TNF (adalimumab, etanercept, infliximab, certolizumab pegol), interleukin (IL)-12/23 (ustekinumab), IL-17A (secukinumab, ixekizumab), IL-17RA (brodalumab) and IL-23p19 (guselkumab, risankizumab, and tildrakizumab) for the primary outcome serious infections, at 10– 16 weeks, 1 year and 3 years. Peto’s method (fixed effect model) was used to estimate the pooled odds ratio (OR) in meta-analyses comparing biologics with one another, methotrexate, or placebo.

**Results:** Forty-three publications (49 trials) consisting of 29,724 participants met our inclusion criteria. Serious infections across all studies were low (n=97) at 10–16 weeks and did not show an increased risk with biologic therapies compared with placebo. Seven head-to-head RCTs were identified, most of which showed no significant difference in the risk of serious infections at 10–16 weeks and at 1 year. Adalimumab was not associated with a significant increased risk of serious infections compared with methotrexate in children at 10–16 weeks.

**Conclusions:** Biologics for psoriasis were not associated with an increased risk of serious infections compared with placebo or one another at 10–16 weeks. Longer-term, real-world data with larger sample sizes are warranted.

## INTRODUCTION

Biologic therapies have transformed the care of patients with moderate-to-severe psoriasis. These highly efficacious medications inhibit the immune pathways implicated in the pathogenesis of psoriasis. There are currently 11 biologic therapies available to patients with psoriasis in the United Kingdom (U.K.) and an emerging challenge is deciding which treatment to select.

Drug safety is an important differentiator of therapies for patients and clinicians, which is different from “perceived” drug safety; in this report, we are addressing the former. Since cytokines targeted by biologics are also implicated in the defence against pathogenic organisms, it is important to understand the potential serious infection risks associated with biologic therapies.

Therefore, we have updated our previously published systematic review and meta-analysis of risk of serious infection to incorporate data on more recently licensed agents and provided part of the evidence used to inform the British Association of Dermatologists (BAD) updated biologic guidelines for psoriasis and to help patients and clinicians choose between the different biologic therapies for psoriasis.^1^ In addition to the five previously included biologic therapies (infliximab, etanercept, adalimumab, ustekinumab and secukinumab), this systematic review sought to incorporate data on risk of serious infections associated with newer biologic therapies targeting TNF (certolizumab pegol), interleukin (IL)-17A (ixekizumab), IL-17RA (brodalumab) and IL-23p19 (guselkumab, risankizumab and tildrakizumab).

## METHODS

### Participants

This systematic review and meta-analysis work was conducted in accordance with the PRISMA statement, and the original protocol was registered on the PROSPERO International prospective register of systematic reviews (2015: CRD42015017538); this has been amended to incorporate data from more recently licensed biologics. Individuals with moderate-to-severe psoriasis receiving biologics primarily for their skin disease were included. Where available, strata included children (up to 12 years) and young people (12-17 years), people with different psoriasis phenotypes (plaque, guttate, pustular, and nail psoriasis), and people receiving a second biologic after treatment failure of the first. Factors such as biologic dosing regimen, methotrexate dose, disease severity, skin type and ethnicity, psoriatic arthritis, and body mass index were examined for subgroup analysis,^2,3^ where heterogeneity was present. Randomized controlled trials (RCTs) and cohort studies (for long-term efficacy/effectiveness or safety data) were eligible for inclusion. Studies were included if they investigated one or more of the following interventions: biologics targeting TNF (adalimumab, etanercept, infliximab, certolizumab pegol), interleukin (IL)-12/23 (ustekinumab), IL-17A (secukinumab, ixekizumab), IL-17RA (brodalumab) and IL-23p19 (guselkumab, risankizumab, and tildrakizumab); the comparison could be any of the listed biologics, methotrexate, or placebo. Conference abstracts were excluded.

Studies were excluded if there were fewer than 50 participants (e.g. fewer than 25 participants per arm). Studies with a population including a proportion greater than 50% being treated primarily for psoriatic arthritis were considered indirect evidence and excluded from this review.

### Search and study selection

The systematic literature search was conducted in PubMed, Medline, Embase and Cochrane electronic databases from inception to 7^th^ September 2018 for the more recently licensed biologics, and from 5^th^ October 2016 to 7^th^ September 2018 for biologics included in the previous systematic review.^4^ Following de-duplication, titles were reviewed by one reviewer (LSE).

Abstracts were screened against the eligibility criteria outlined in the protocol by two independent reviewers (ZZNY and Zarif K. Jabbar-Lopez); any disagreements were resolved by an arbitrator (CHS). Full texts were obtained and assessed for inclusion against the eligibility criteria outlined in the protocol (LSE). Reference lists of systematic reviews and meta-analyses were screened for additional studies not identified by the systematic literature search (LSE). The included full-text articles were distributed amongst the authors (LM, MCE, MFMM) for critical appraisal and data extraction using a standardized data extraction tool (RevMan version 5.3., The Nordic Cochrane Centre, The Cochrane Collaboration, 2014, Copenhagen) and confirmed by another (LSE).

### Outcomes of interest

The primary outcome was serious infections (investigator-defined) at 10–16 weeks, 1 year (±4 weeks), and 3 years in accordance with our systematic review protocol (Table S1; see supporting information) underpinning the updated BAD guidelines for biologic therapy for psoriasis.^1^

### Meta-analysis and quality assessment

Included studies reporting serious infections data were pooled in a meta-analysis using RevMan 5.3, visualised via forest plots. Peto’s method with a fixed effect model was used to estimate the pooled odds ratio (OR) as it has been shown to give the least biased results for rarer events.^5^ Statistical heterogeneity was assessed using the *I*^*2*^ test. Critical appraisal was conducted using a checklist from the National Institute for Health and Care Excellence (NICE) which assessed selection bias, performance bias, attrition bias, measurement bias, and outcome-reporting bias. Grading of Recommendations Assessment, Development and Evaluation (GRADE) criteria were used to assess the certainty of evidence for outcomes across studies using software from the GRADE Working Group (GRADEpro Guideline Development Tool [Software]. McMaster University, 2015 (developed by Evidence Prime, Inc.); available from www.gradepro.org). The certainty of evidence for outcomes across studies was classed as *very low, low, moderate*, and *high*, based on assessment of the risk of bias, inconsistency, indirectness, and publication/reporting bias. In alignment with GRADE criteria, observational studies were automatically downgraded prior to assessment and rated as low-certainty evidence. Observational studies were graded upwards if the magnitude of the treatment effect was large, if there was evidence of a dose-response relationship, and if all plausible confounders would have ordinarily decreased the magnitude of an apparent treatment effect. Sensitivity analyses were performed using the Mantel-Haenszel risk ratios to add robustness to the results.

## RESULTS

The systematic literature search yielded 3727 publications; following de-duplication, 2491 were screened for eligibility (Figure 1). A total of 114 full-text articles were assessed for inclusion against the eligibility criteria outlined in the protocol (Table S1; see supporting information), resulting in the exclusion of a further 71 articles. The reasons for exclusion are presented in Table S3 (see supporting information). Therefore, a total of 43 publications (49 trials) consisting of 29,724 participants from RCTs met the inclusion criteria and were included in the systematic review (Table S2; see supporting information). Twenty-four RCTs excluded patients with a history of serious infections and 28 RCTs included patients who had previously received biologic therapy.

**Figure 1.**
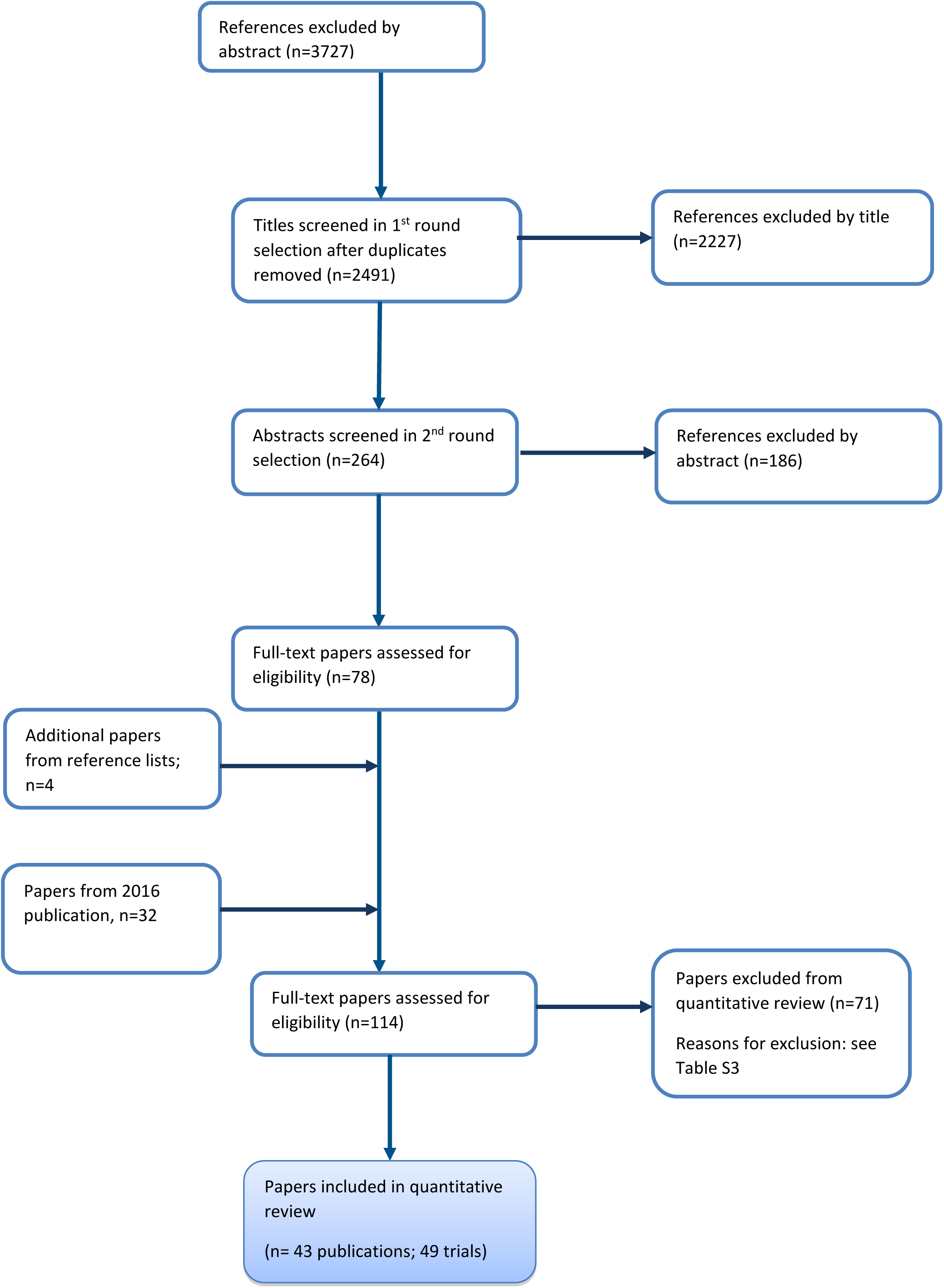
Flow diagram showing the identification of literature in the PRISMA statement format

A dose-independent analysis was conducted. There was no statistically significant heterogeneity indicated for any of the comparisons, therefore, it was not necessary to conduct any subgroup analysis based on different dosing regimens.

In total, the number of serious infections reported across all the included RCTs at 10–16 weeks was low (n=97) and 11 of the included studies did not report any serious infection in any intervention arm. Most of the included studies reported data at the primary efficacy time points of 10–16 weeks, with limited data available at 1 year and none at 3 years.

### Risk of bias

The majority of studies (29/43, 67%) were rated as having a low, overall risk of bias (5/43 (12%) rated as high risk, 7/43 (16%) rated as very high risk). Similarly, 35/43 (81%) and 33/43 (77%) studies were rated as having a low risk of selection and performance bias, respectively. Generally, the risk of attrition bias was low across all studies. All included studies were supported financially by industry except for one which did not report the source of funding.^6^

The overall quality/certainty of the evidence for each outcome was assessed using GRADE criteria, and most of the studies were rated as low or very low certainty. One study comparing risankizumab with ustekinumab at 1 year had a rating of moderate certainty;^7^ studies comparing tildrakizumab with placebo^8,9^ and risankizumab with ustekinumab (UltIMMa-1 and UltIMMa-2)^10^ were rated as high certainty. The main reasons for downgrading the certainty of evidence were due to imprecision and high risk of biases. Sensitivity analyses using Mantel-Haenszel methods for fixed- and random-effects models did not influence the conclusions for the biologic vs. placebo comparisons at 10–16 weeks (Figures S1 and S2; see supporting information).

### Evidence from RCTs: serious infections with biologic therapies compared with placebo in adults

#### At 10–16 weeks

Fifteen studies (involving 20 trials) investigating different biologic therapies were considered eligible for inclusion, in addition to the 22 studies identified in the previous systematic review. There was no increased risk of infection associated with any of the biologic agents compared with placebo (Figure 2; Table S4 – see supporting information). Heterogeneity was not found across the different biologic therapies except for etanercept which had moderate heterogeneity; however, the impact of this heterogeneity was minimal due to the non-statistically significant *I*^*2*^ value (p=0.10) and the small pooled Peto OR (Figure 2; Table S4 – see supporting information).

**Figure 2.**
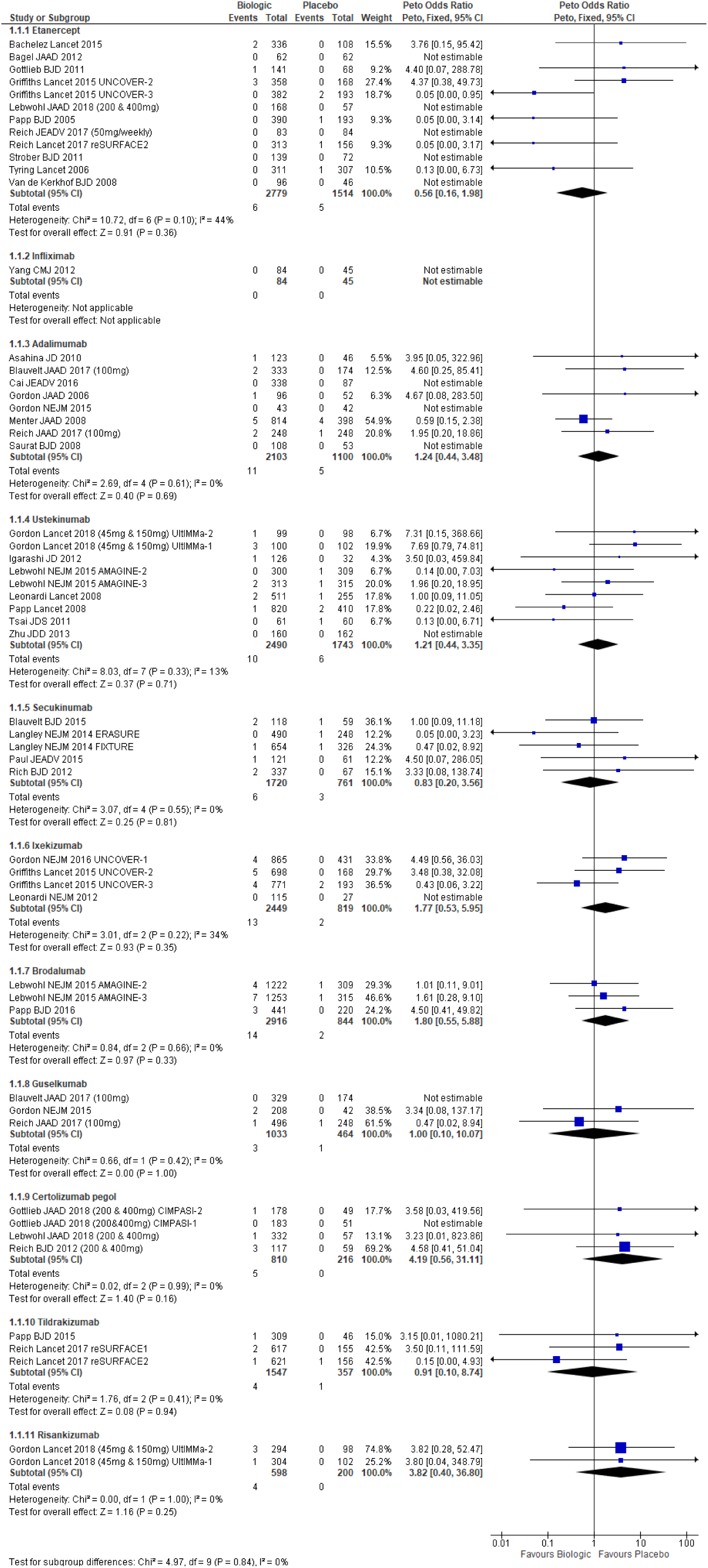
Dose-independent comparison for biologic vs. placebo, serious infections at 10–16 weeks

Studies for the newer biologics were conducted in centres worldwide, the majority of which were in North America and Europe, including studies which recruited Asian populations from countries such as Japan, South Korea, Taiwan, and China.

Assessment using the GRADE criteria showed that the certainty of the evidence for each outcome varied across the different biologics, however, the majority were rated as low or very low certainty, mainly due to imprecision. The certainty of evidence was rated as high for studies investigating tildrakizumab, low for studies investigating etanercept, secukinumab, ixekizumab, brodalumab, guselkumab, certolizumab pegol and risankizumab, and very low for studies investigating adalimumab and ustekinumab. The overall certainty of evidence could not be assessed for one study comparing infliximab with placebo due to a lack of events in both treatment arms.

### Evidence from RCTs: serious infections with biologic therapies compared with methotrexate in children and adolescence

#### At 10–16 weeks

In the previous systematic review, one study was identified investigating adalimumab versus methotrexate in adults.^11^ This systematic review identified one additional study investigating adalimumab (n=77) versus methotrexate (n=37) in children and adolescents (aged ≥4 to <18 years).^12^ Only one event was reported in the adalimumab arm, with no events in the methotrexate arm; the Peto OR showed a non-statistically significant increased risk of serious infections (Figure 3; Table S4 – see supporting information). The very large confidence interval is indicative of the small sample size and lack of events in a treatment arm.

**Figure 3:**
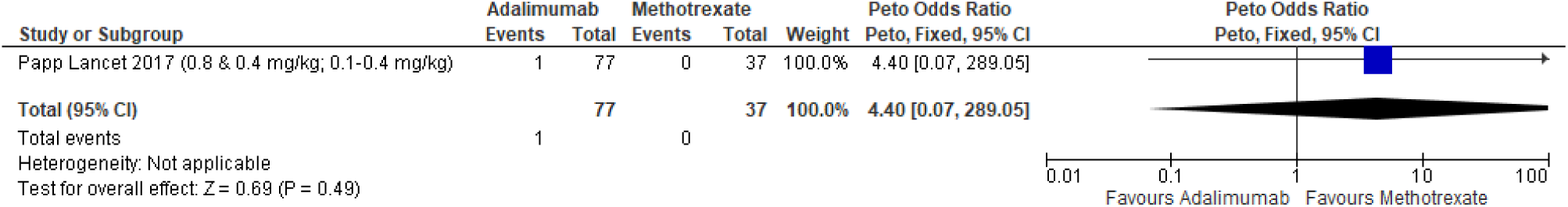
Dose-independent comparison for adalimumab vs. methotrexate, serious infections at 10–16 weeks in ≥4 to <18 years

### Evidence from RCTs: serious infections between biologic therapies in adults

#### At 10–16 weeks

Seven additional studies (nine trials) were considered eligible for inclusion. One study reported results from two trials (AMAGINE-2 and AMAGINE-3) comparing brodalumab with ustekinumab; the pooled Peto OR showed no statistically significant difference in risk of serious infections (Figure 4; Table S4 – see supporting information). Another study which reported results from two trials (UltIMMa-1 and UltIMMa-2) compared risankizumab with ustekinumab; the pooled Peto OR showed a non-statistically significant decrease in the risk of serious infections with risankizumab (Figure 5; Table S4 – see supporting information). Three studies comparing guselkumab with adalimumab also showed a non-statistically significant decrease in the risk of serious infections with guselkumab (Figure 6; Table S4 – see supporting information). The Peto OR showed no statistically significant increased or decreased risk of serious infections with certolizumab pegol (Figure 7; Table S4 – see supporting information) or tildrakizumab (Figure 8; Table S4 – see supporting information) when compared with etanercept. These results should be interpreted with caution since all comparisons had wide confidence intervals which is likely due to the small number or lack of events in any treatment arm, and the small number of studies combined for pooled estimates.

**Figure 4:**
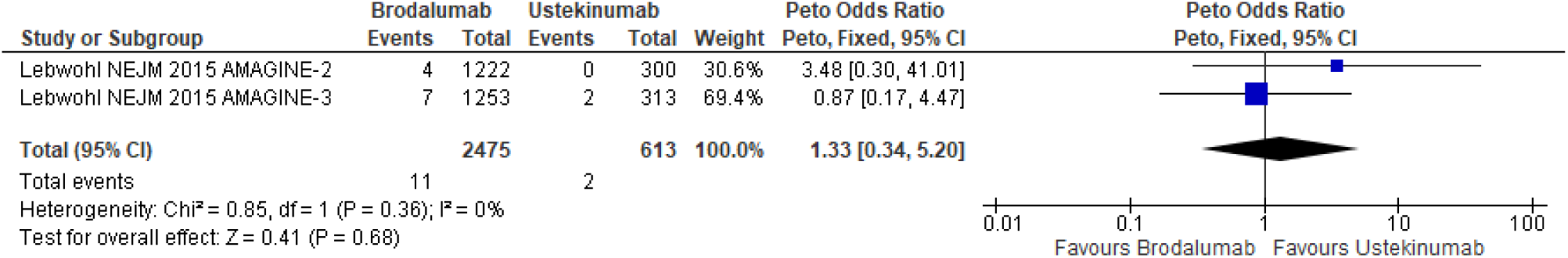
Dose-independent comparison for brodalumab vs. ustekinumab, serious infections at 10–16 weeks

**Figure 5:**
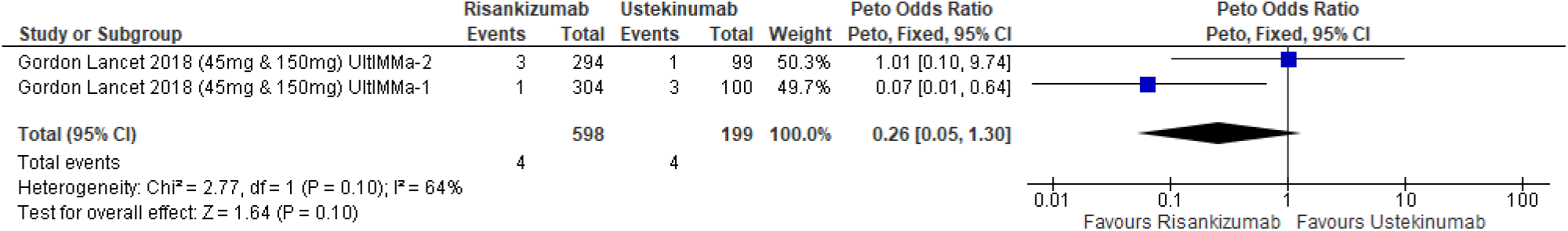
Dose-independent comparison for risankizumab vs. ustekinumab, serious infections at 10–16 weeks

**Figure 6:**
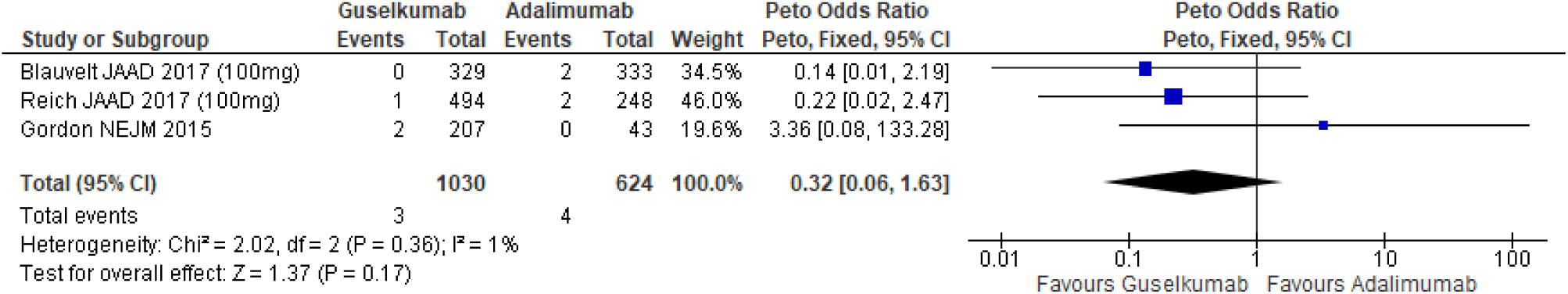
Dose-independent comparison for guselkumab vs. adalimumab, serious infections at 10– 16 weeks

**Figure 7:**
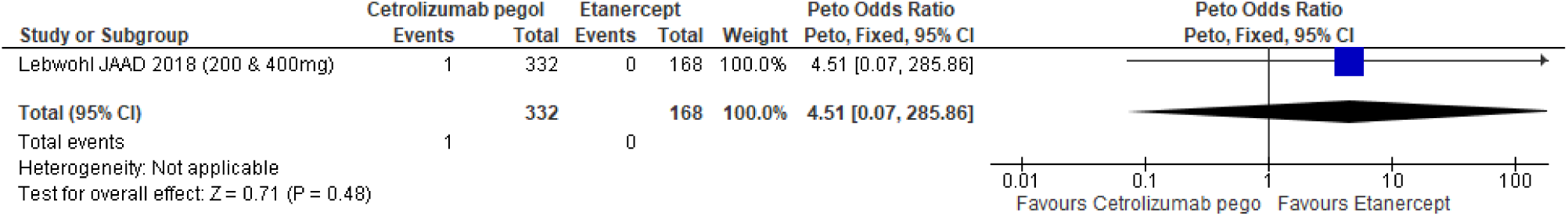
Dose-independent comparison of certolizumab pegol vs. etanercept, serious infections at 10–16 weeks

**Figure 8:**
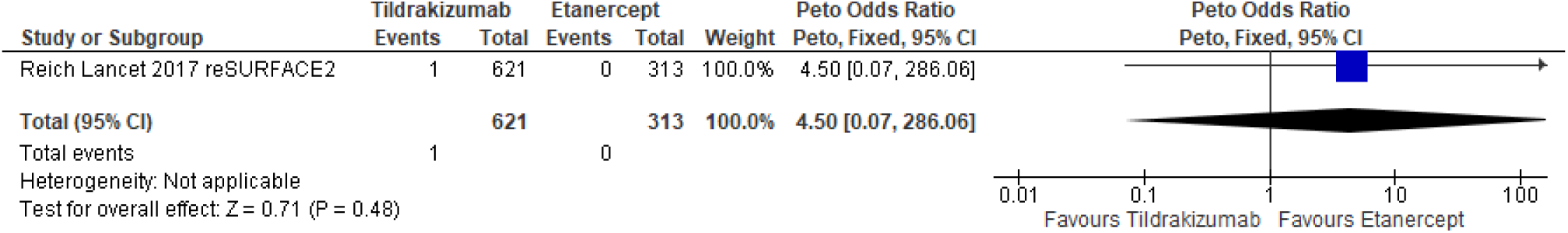
Dose-independent comparison of tildrakizumab vs. etanercept, serious infections at 10– 16 weeks

For the GRADE assessment, the certainty of evidence was generally low or very low (Tables S7– S11; see supporting information), except for one study comparing risankizumab with ustekinumab^10^ which was rated as high (Table S9; see supporting information).

#### At 1 year

Two additional studies were considered eligible for inclusion.^7,13^ There was no difference in risk of serious infections between guselkumab and adalimumab (Figure 9; Table S5 – see supporting information),^13^ and a reduced risk of serious infections associated with risankizumab compared with ustekinumab (Figure 10; Table S4 – see supporting information; NNTH 15, 95% CI 62.43–6.66).^7^

**Figure 9:**
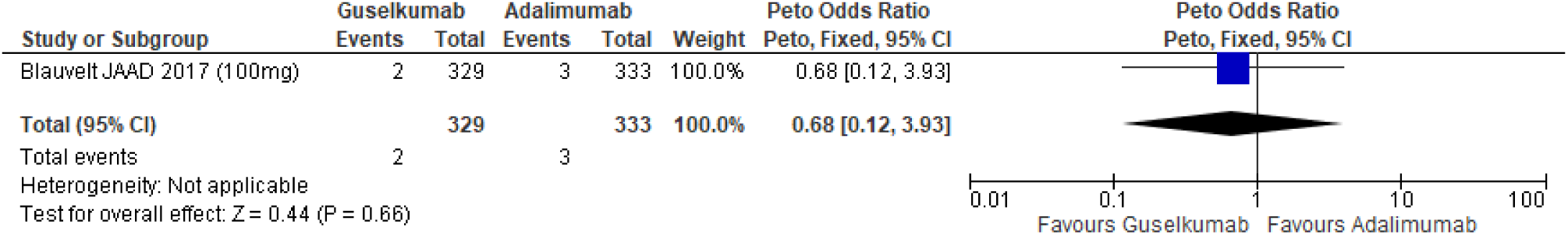
Dose*-*independent comparison for guselkumab vs. adalimumab, serious infections at 1 year

**Figure 10:**
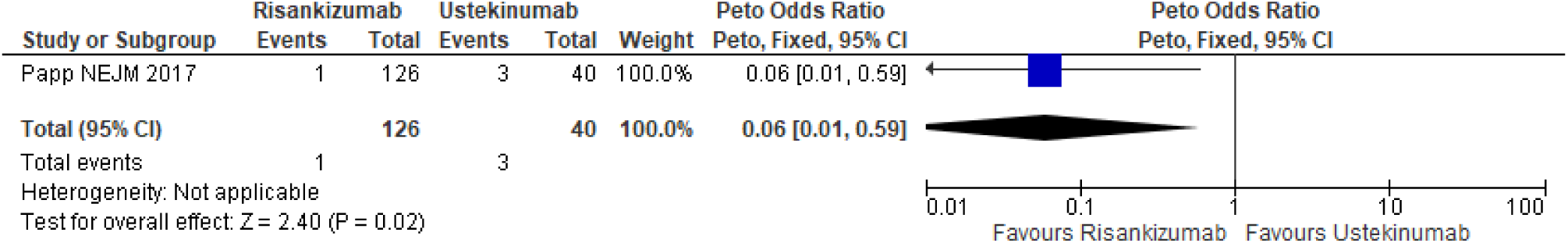
Dose-independent comparison for risankizumab vs. ustekinumab, serious infections at 1 year

## DISCUSSION

In this updated systemic review, we find that none of the currently licensed biologic therapies used for the treatment of psoriasis, including the newer biologics (ixekizumab, brodalumab, guselkumab, certolizumab pegol, tildrakizumab and risankizumab)^4^ had a higher risk of serious infections compared with placebo with short-term treatment, with low event rates across all treatment arms. When considering differential risk of infection between biologics, in one of seven head-to-head comparator studies identified, risankizumab had a reported reduced risk of serious infection (OR = 0.06, 95% CI 0.01–0.59) compared with ustekinumab at one year. However, the event rates from this single study were very low, and so this difference needs to be interpreted with caution and may not be either generalizable or clinically relevant.

### Findings in context and clinical relevance

A focussed review investigating IL-17 and IL-23 antagonists in psoriasis and psoriatic arthritis populations reported low event rates at 16 and 52 weeks with no increased risk of serious infection compared with placebo;^14^ these results are reassuring. However, when considering these findings in the context of clinical practice there are important limitations. Firstly, because the event rates are low (and trials are powered for efficacy not safety), the confidence intervals are large, and thus imprecision around the estimates. Secondly, these data only relate to short term use, with the longest time point of 1-year being reported by only two studies.^7,13^ Thirdly, findings lack generalizability due to the homogenous patient populations in the trials which are not reflective of real-world settings. Finally, there were few with few head-to-head comparisons, or biologics compared to methotrexate, so risk estimates on clinically relevant comparisons are missing.

Long-term, real-world data identified in our search but not fulfilling our inclusion criteria, as well as more recent data, are consistent with this reassuring picture. A recent investigation in patients with psoriasis (n=5617) using prospective national registry data from the BAD Biologic Interventions Register (BADBIR) that did not meet our inclusion criteria has shown no statistically significant increased risk of serious infections with etanercept, adalimumab and ustekinumab compared with conventional, non-biologic systemic therapies.^15^ A second, retrospective, single-centre cohort study that did not meet our inclusion criteria^16^ concluded that no statistically significant difference was detected for the risk of serious infections when comparing people with psoriasis and psoriatic arthritis exposed to biologic and nonbiologic therapies. The findings from these real-world studies are consistent with ours. Since our search, a retrospective cohort study provides real-world evidence of the risk of serious infections in adults in the USA (diagnosed with psoriasis or psoriatic arthritis between 2015 and 2018) treated with IL-17, IL-12/23, or TNF inhibitors amongst biologic-naïve and experienced biologic patients.^17^ The risk of serious infections was similar amongst the new users of IL-17 and TNF inhibitors, whilst the persons newly treated with an IL-12/23 were less likely to be hospitalized. However, this study had a relatively short follow-up period of 6 months. Data on infliximab remains sparse although in a UK BADBIR cohort study was associated with a two-fold increased risk of serious infections compared with conventional non-biologic systemic treatment (adjusted HR 1.95, 95% CI 1.01–3.75).^18^

### Limitations and future directions

We established that it was not possible to calculate a pooled Peto OR for all biologic therapies versus placebo due to the inclusion of three-arm trials, each investigating two different biologics and placebo (UltIMMa-1, UltIMMa-2, AMAGINE-2, AMAGINE-3, UNCOVER-2, UNCOVER 3, reSURFACE-2); doing so would have meant double-counting the number of patients in the placebo arms, which would create a unit-of-analysis error,^19^ and would have underestimated the overall effect of the biologic therapies. It was not possible to combine treatment groups to create a single, pair-wise comparison, as we did not think it was appropriate to combine patients in the placebo group with those in a biologic therapy group. One way to overcome this issue would be to include serious infection in updating our network meta-analysis;^20^ we did not conduct such analysis as this is an update of our previous systematic review.^4^

Our systematic review included a limited number of head-to-head trials and those comparing biologics to conventional systemic therapies. Moreover, the studies included in the original systematic review did not report their definition for the outcome of serious infections adequately, and this remained a limitation with the additional studies identified in this update. The terminology was ambiguous and not consistent amongst the included studies. We suggest that the term “serious infection” be used and formally defined as a serious adverse event that is an infection as per the International Conference on Harmonisation (ICH) guideline.^21^ The trial report should also state the criteria to which a “medically important” infection is defined by the investigators.

Our previous systematic review identified only two studies conducted in children, both did not report any serious infection in the treatment and placebo arms.^4^ The additional study identified in this systematic review,^12^ evaluating 114 patients only, reported one event in the adalimumab arm. There is a need for trials conducted in children and adolescent populations with psoriasis assessing the risk of serious infections between different biologics or compared with conventional systemic therapies.

## CONCLUSION

This updated systematic review shows no significant increased risk of serious infection with biologic therapies for psoriasis when compared to placebo, or in limited comparisons with each other during the primary efficacy time points of 10–16 weeks. These data are reassuring. Generalisability of these findings, particularly for the newer classes of biologic therapies, require real-world observational data from population-based datasets, for example from large-scale registries and well curated, healthcare records.

## Data Availability

N/A

## ACKNOWLEDGEMENTS

We would like to acknowledge Zarif K. Jabbar-Lopez for his contribution at the early stages. This systematic review and meta-analysis were supported by the BAD to inform the updated guidelines for biologic interventions for psoriasis. We

