## Supporting information for "Risk of Serious Infections in Patients with Psoriasis on Biologic Therapies: An Updated Systematic Review and Meta-Analysis"

**Table S1. Protocol for systematic review**

| | |
|---|---|
| **Review question** | In people with psoriasis (all types), what is the risk of serious infection of biologics (adalimumab, brodalumab, certolizumab pegol, etanercept, guselkumab, infliximab, ixekizumab, risankizumab, secukinumab, tildrakizumab or ustekinumab) compared with each other, with methotrexate or with placebo? |
| **Objectives** | The aim of this review is to assess the risk of serious infection of biologics (adalimumab, brodalumab, certolizumab pegol, etanercept, guselkumab, infliximab, ixekizumab, risankizumab, secukinumab, tildrakizumab or ustekinumab) compared with each other, with methotrexate, and with placebo (or no treatment). |
| **Population** | All people with psoriasis with moderate to severe disease[1]being treated primarily for their skin disease |
| **Strata** | The following groups will be considered separately if data are available:<br>• Children (up to 12 yrs) & young people (12-18 yrs)<br>• Different psoriasis phenotypes – i.e. plaque, guttate, pustular (generalized pustular psoriasis, localized forms i.e. palmoplantar pustulosis and acrodermatitis continua of Hallopeau) and nail psoriasis<br>• People receiving a second biologic (after the failure of the first) |
| **Subgroups** | The following factors will be considered for subgroup analysis if heterogeneity is present:<br>• Methotrexate dose<br>• Biologics dose (NICE-approved vs. non-NICE approved dose)<br>• Disease severity (moderate to severe vs. very severe)<br>• Skin type (Fitzpatrick scale) and ethnicity [Safety only]<br>• Psoriatic arthritis<br>• BMI/body weight |
| **Intervention** | • Adalimumab<br>• Brodalumab<br>• Certolizumab pegol<br>• Etanercept<br>• Guselkumab<br>• Infliximab<br>• Ixekizumab<br>• Risankizumab<br>• Secukinumab<br>• Tildrakizumab<br>• Ustekinumab (2 doses based on body weight)<br>Note: all doses and durations will be included; sensitivity analyses for licensed doses only will be carried out in the network meta-analyses |
| **Comparison** | • Placebo<br>• Adalimumab<br>• Brodalumab<br>• Certolizumab pegol<br>• Etanercept |

[1] Defined as requiring systemic therapy and/or PASI or BSA>10 (CPP) and/or PGA of at least moderate

| | |
|---|---|
| | • Guselkumab<br>• Infliximab<br>• Ixekizumab<br>• Risankizumab<br>• Secukinumab<br>• Tildrakizumab<br>• Ustekinumab<br>• Methotrexate (within standard dose range 15-25 mg) |
| **Outcomes** | Outcome to be extracted at 10 – 16 weeks,[2] 1 year (±4 weeks), and 3 years:<br>• Serious infection and TB |
| **Study design** | • RCTs or systematic reviews<br>• Cohort studies for long-term data |
| **Population size and directness** | • Sample size >50 (i.e. 25 in each arm)<br>• Studies with indirect populations will not be considered<br>• Studies in populations where the proportion being treated primarily for psoriatic arthritis was greater than 50% will be considered indirect |
| **Setting** | • Secondary care<br>• Tertiary care<br>• Community settings in which NHS care is received |
| **Search Strategy** | See section S1.1 |
| **Review strategy** | Appraisal of methodological quality<br>• The methodological quality of each study will be assessed using NICE checklists and the synthesis of data |

### Section S1.1. Search strategy

Searches for clinical reviews we (ProQuest/Dialog), the Cochrane Library (Wiley). Typically, searches were constructed in the following way:
Searches for **clinical reviews** were run in Medline (ProQuest/Dialog),
Embase (ProQuest/Dialog), the Cochrane Library (Wiley). Typically, searches were constructed in the following way:

A PICO format was used for intervention searches. **Population** (P) terms were combined with **Intervention** (I) and s ometimes **Comparison** (C) terms (as indicated in the tables under each individual question in Section S.1.4). An inte rvention can be a drug, a procedure, or a diagnostic test. **Outcomes** (O) are rarely used in search strategies for inter ventions. Study type filters were added where appropriate (see S.1.2).

Where possible searches were limited to papers published in English.

| **Section S1.2** | **Study filter terms** |
|---|---|
| S1.2.1 | Systematic reviews (SR) |
| S1.2.2 | Randomized controlled trials (RCT) |
| S1.2.3 | Observational studies |
| **Section S1.3** | **Standard population search strategy**<br>This population was used for all search questions unless stated |
| **Section S1.4** | **Searches for specific questions** with intervention (and population where different from S.1.2) |
| S1.4.1 | Efficacy and safety in biologic therapy (original biologics) |
| S1.4.2 | Efficacy and safety in biologic therapy (new biologics) |

[2] In line with current NICE technology appraisals

### Section S1.2. Search terms for types of studies involved

#### S1.2.1 Systematic reviews

**Medline and EMBASE search terms**

| | |
|---|---|
| 1. | review[*1] |
| 2. | AB,TI(systematic[*4] OR evidence[*2] OR methodol[*6] OR quantitativ[*2] OR analys[*2] OR assessment[*2]) |
| 3. | 1 and 2 |
| 4 | S2 AND (dtype("review")) |
| 5 | (systematic pre/0 review[*1]) |
| 6. | (meta-analys[*2]) |
| 7. | (dtype("meta-analysis")) |
| 8. | AB,TI(meta-analy* or metanaly* or metaanaly* or meta pre/0 analy*) |
| 9. | AB,TI((systematic[*4] or evidence[*2] OR methodol[*6] OR quantitativ[*2] ) n/5 (review[*1] or survey[*1] or overview[*1])) |
| 10. | AB,TI((pool* or combined or combining) n/2 (data or trial[*1] or studies or results)) |
| 11. | 3 OR 4 OR 5 OR 6 OR 7 OR 8 OR 9 OR 10 |

**PubMed Search terms**

| | |
|---|---|
| 1. | review |
| 2. | systematic*[Title/Abstract] OR evidence*[Title/Abstract] OR methodol*[Title/Abstract] OR quantitativ*[Title/Abstract] OR analys*[Title/Abstract] OR assessment*[Title/Abstract] |
| 3. | 1 AND 2 |
| 4 | "systematic review" |
| 5 | "meta-analysis"[Publication Type] OR "meta-analysis as topic"[MeSH Terms] (meta-analys*) OR (meta-analy*[Title/Abstract] OR metanaly*[Title/Abstract] OR metaanaly*[Title/Abstract] OR meta analy*[Title/Abstract]) |
| 6. | (systematic*[Title/Abstract] OR evidence*[Title/Abstract] OR methodol*[Title/Abstract] OR quantitative*[Title/Abstract]) AND (review*[Title/Abstract] OR survey*[Title/Abstract] OR overview*[Title/Abstract]) |
| 7. | (pool*[Title/Abstract] OR combined[Title/Abstract] OR combining[Title/Abstract]) AND (data[Title/Abstract] OR trials[Title/Abstract] OR studies[Title/Abstract] OR results[Title/Abstract]) |
| 8. | 3 OR 4 OR 5 OR 6 OR 7 |

#### S1.2.2 Randomised controlled trial (RCT)

**Medline and EMBASE search terms**

| | |
|---|---|
| 1. | (randomi$3 PRE/0 control$3 PRE/0 trial$1) OR (control$3 PRE/0 clinical PRE/0 trial$1) |
| 2. | DTYPE("randomized controlled trial") OR DTYPE("controlled clinical trial") |
| 3. | AB("randomized" OR "randomised") OR AB("placebo") OR AB("randomly") |
| 4. | EMB.EXACT("crossover procedure") OR EMB.EXACT("double blind procedure") OR EMB.EXACT("single blind procedure") OR (EMB.EXACT("randomized controlled trial") OR EMB.EXACT("randomized controlled trial (topic)" |
| 5. | mjmesh.exact("Clinical Trials as Topic") |
| 6. | AB,TI("crossover$2" OR "(cross PRE/0 over$2)" OR "cross-over$2") OR AB,TI((doubl[*3] OR singl[*3]) NEAR/1 blind[*4]) OR AB,TI("assign$5" OR "allocat$4" OR "volunteer$3") |
| 7. | 1 OR 2 OR 3 OR 4 OR 5 OR 6 |

**PubMed search terms**

| | |
|---|---|
| 1. | (randomized controlled trials as topic[MeSH Terms]) OR controlled clinical trials as topic[MeSH Terms] OR (randomi* controlled trial* OR randomi* control trial* OR RCT* OR non-randomi* controlled trial* OR non-randomi* control trial* OR controlled clinical trial*) |
| 2. | randomized[Title/Abstract] OR randomised[Title/Abstract] OR randomly[Title/Abstract] OR placebo[Title/Abstract] OR trial[Title] |
| 3. | crossover*[Title/Abstract] OR cross over*[Title/Abstract] OR cross-over*[Title/Abstract] |
| 4. | (doubl*[Title/Abstract] OR singl*[Title/Abstract]) AND (blind[Title/Abstract] OR blind*[Title/Abstract]) |
| 5. | 1 OR 2 OR 3 OR 4 |

#### S1.2.3 Observational studies

**Medline and EMBASE search terms**

| | |
|---|---|
| 1. | MESH.EXACT.EXPLODE("Clinical Trial") OR MESH.EXACT.EXPLODE("Clinical Trials as Topic") OR EMB.EXACT.EXPLODE("clinical trial (topic)") OR EMB.EXACT.EXPLODE("clinical trial") |
| 2. | EMB.EXACT("controlled study") OR (controlled PRE/0 stud$3) |
| 3. | EMB.EXACT.EXPLODE("evaluation study") OR MESH.EXACT.EXPLODE("Evaluation Studies") OR EMB.EXACT.EXPLODE("prospective study") OR MESH.EXACT.EXPLODE("Prospective Studies") OR MESH.EXACT.EXPLODE("Follow-Up Studies") OR MESH.EXACT.EXPLODE("Epidemiologic Studies") OR EMB.EXACT.EXPLODE("longitudinal study") OR MESH.EXACT.EXPLODE("Longitudinal Studies") OR EMB.EXACT.EXPLODE("cohort analysis") |
| 4. | AB,TI(cohort PRE/0 stud$3) |
| 5. | AB,TI("crossover$2" OR "(cross PRE/2 over$2)" OR "cross-over$2") NEAR/2 AB,TI("design$3" OR "stud$3" OR "procedure$1" OR "trial$3") |
| 6. | EMB.EXACT("crossover procedure") OR MESH.EXACT("Cross-Over Studies") |
| 7. | 1 OR 2 OR 3 OR 4 OR 5 OR 6 |

**PubMed search terms**

| | |
|---|---|
| 1. | "clinical trial"[Publication Type] OR "clinical trials as topic"[MeSH Terms] OR "clinical trial"[All Fields] |
| 2. | "evaluation studies"[Publication Type] OR "evaluation studies as topic"[MeSH Terms] OR "evaluation studies"[All Fields] |
| 3. | "follow-up studies"[MeSH Terms] OR ("follow-up"[All Fields] AND "studies"[All Fields]) OR "follow-up studies"[All Fields] OR "follow up studies"[All Fields] |
| 4. | "prospective studies"[MeSH Terms] OR ("prospective"[All Fields] AND "studies"[All Fields]) OR "prospective studies"[All Fields] |
| 5. | "epidemiologic studies"[MeSH Terms] OR "epidemiologic studies"[All Fields] OR "epidemiological studies"[All Fields] |
| 6. | cohort studies[MeSH Terms]) OR (cohort study[Title/Abstract] OR cohort studies[Title/Abstract] |
| 7. | (crossover*[Title/Abstract] OR cross over*[Title/Abstract] OR cross-over*[Title/Abstract]) AND (design*[Title/Abstract] OR study*[Title/Abstract] OR studies*[Title/Abstract] OR procedure*[Title/Abstract] OR trial*[Title/Abstract]) |
| 8. | 1 OR 2 OR 3 OR 4 OR 5 OR 6 OR 7 |

### Section S1.3. Standard population search strategy

**MEDLINE and EMBASE search terms**

| | |
|---|---|
| 1. | EMB.EXACT.EXPLODE("psoriasis") OR mesh.exact("Psoriasis") OR AB,TI,IF("psoria*") |
| 2. | AB,TI,IF("pustulo*" n/3 "palm*") |

| | |
|---|---|
| 3. | S1 OR S2 |
| 4. | EMB.EXACT("letter") OR letter[*1] OR DTYPE("letter") |
| 5. | DTYPE("editorial") OR DTYPE("historical article") OR DTYPE("anecdote") OR DTYPE("note") OR DTYPE("commentary") |
| 6. | EMB.EXACT("case report") OR (case PRE/0 report$1) OR DTYPE(case report$1) |
| 7. | EMB.EXACT("case study") OR (case PRE/0 stud[*3]) OR DTYPE(case study) OR AB,TI(case PRE/0 control$1) |
| 8. | (EMB.EXACT.EXPLODE("animal") OR MESH.EXACT.EXPLODE("Animals")) AND (animal(yes)) |
| 9. | EMB.EXACT("nonhuman") |
| 10. | EMB.EXACT.EXPLODE("animal experiment") OR EMB.EXACT.EXPLODE("experimental animal") OR EMB.EXACT.EXPLODE("animal model") OR MESH.EXACT.EXPLODE("Animal Experimentation") OR MESH.EXACT.EXPLODE("Animals, Laboratory") OR MESH.EXACT.EXPLODE("Models, Animal") |
| 11. | MESH.EXACT.EXPLODE("Rodentia") OR EMB.EXACT.EXPLODE("rodent") |
| 12. | S4 OR S5 OR S6 OR S7 OR S8 OR S9 OR S10 OR S11 |
| 13. | S3 NOT S12 |

**PubMed search terms**

| | |
|---|---|
| 1. | "psoriasis"[MeSH Terms] OR "psoriasis"[All Fields] OR psoria*[Title/Abstract] |
| 2. | (pustulo*[Title/Abstract]) AND (palmopl*[Title/Abstract] OR palmari*[Title/Abstract] OR palmar[Title/Abstract]) |
| 3. | 1 OR 2 |
| 4. | "letter"[Publication Type] OR "correspondence as topic"[MeSH Terms] OR "letter"[All Fields] OR “letter*”[All fields] |
| 5. | "editorial"[Publication Type] OR "historical article"[Publication Type] OR "comment"[Publication Type] |
| 6. | "case reports"[Publication Type] OR case report* OR "case study"[All Fields] OR "case studies"[All Fields] OR case control stud*[Title/Abstract] |
| 7. | animal Filters: Other Animals |
| 8. | nonhuman[All Fields] |
| 9. | "animals, laboratory"[MeSH Terms] OR "laboratory animals"[All Fields] OR "experimental animal"[All Fields] OR "animal experimentation"[MeSH Terms] OR "animal experimentation"[All Fields] OR "animals, laboratory"[MeSH Terms] OR "laboratory animals"[All Fields] OR "laboratory animal"[All Fields] OR "models, animal"[MeSH Terms] OR "animal models"[All Fields] OR "animal model"[All Fields] OR animal modeling[All Fields] OR animal modelling[All Fields] |
| 10. | "rodentia"[MeSH Terms] OR "rodentia"[All Fields] OR "rodent"[All Fields] OR "rodents"[All Fields] |
| 11. | 4 OR 5 OR 6 OR 7 OR 8 OR 9 OR 10 |
| 12. | 3 NOT 11 |

**Cochrane search terms**

| | |
|---|---|
| 1. | psoria*:ti,ab,kw |
| 2. | pustulo* near/3 palm*:ti,ab,kw |
| 3. | #1 OR #2 |

#### S1.4 Searches for specific questions

**In people with psoriasis (all types), what is the risk of serious infection of biologics (adalimumab, etanercept, infliximab, ixekizumab, secukinumab or ustekinumab) compared with each other, with methotrexate or with placebo?**

Search constructed by combining the columns in the following table using the **and** Boolean operator

| Population | Intervention | Comparison | Study filter used | Date parameters |
|---|---|---|---|---|
| Psoriasis | Biologic therapy (original biologics) | | RCTs, SRs and Observational studies [Medline and EMBASE only] | 05/10/2016 – 07/09/2018 |

##### S.1.4.1 Biologic therapy (original biologics)

**Medline and EMBASE search terms**

| | |
|---|---|
| 1. | (EMB.EXACT("etanercept") OR EMB.EXACT("infliximab") OR EMB.EXACT("adalimumab") OR EMB.EXACT("ustekinumab") OR EMB.EXACT("secukinumab") OR (EMB.EXACT("ixekizumab")) OR AB,TI,IF(etanercept OR infliximab OR adalimumab OR ustekinumab OR secukinumab OR ixekizumab) |
| 2. | AB,TI(cosentyx OR enbrel OR humira OR remicade OR stelara OR taltz ) |
| 3. | MESH.EXACT("Biosimilar Pharmaceuticals") OR EMB.EXACT("biosimilar agent") OR AB,TI("biosimilar$1") OR AB,TI(erelzi OR benepali OR brenzys OR inflectra OR remsima OR flixabi OR ixifi OR zessly OR amgevita OR a mjevita OR solymbic OR cyltezo OR imraldi OR halimatoz OR GP2017) |
| 4. | EMB.EXACT.EXPLODE("monoclonal antibody") OR MESH.EXACT.EXPLODE("Antibodies, Monoclonal") |
| 5. | MESH.EXACT.EXPLODE("Receptors, Tumor Necrosis Factor") OR EMB.EXACT.EXPLODE("tumor necrosis factor receptor") |
| 6. | (EMB.EXACT.EXPLODE("interleukin 12") OR EMB.EXACT.EXPLODE("interleukin 17") OR EMB.EXACT.EXPLODE("interleukin 23")) OR (MESH.EXACT.EXPLODE("Interleukin-12") OR MESH.EXACT.EXPLODE("Interleukin-17") OR MESH.EXACT.EXPLODE("Interleukin-23") OR MESH.EXACT("Interleukins")) |
| 7. | AB,TI(TNF NEAR/1 (antagonis[*3] OR inhibit[*3])) |
| 8. | AB,TI("T cell helper") |
| 9. | AB,TI("anti-TNF") |
| 10. | 1 OR 2 OR 3 OR 4 OR 5 OR 6 OR 7 OR 8 OR 9 OR 10 |

**PubMed search terms**

| | |
|---|---|
| 1. | adalimumab OR etanercept OR infliximab OR ixekizumab OR secukinumab OR ustekinumab OR cosentyx OR enbrel OR humira OR remicade OR stelara OR taltz |
| 2. | biosimilar[Title/Abstract] OR biosimilars[Title/Abstract] OR erelzi OR benepali OR brenzys OR inflectra OR remsima OR flixabi OR ixifi OR zessly OR amgevita OR amje vita OR solymbic OR cyltezo OR imraldi OR halimatoz OR GP2017 |
| 3. | "antibodies, monoclonal"[MeSH Terms] OR "monoclonal antibodies"[All Fields] |
| 4. | "interleukins"[MeSH Terms] OR "interleukins"[All Fields] OR "interleukin-12"[MeSH Terms] OR "interleukin-12"[All Fields] OR "interleukin 12"[All Fields] OR "interleukin-27"[MeSH Terms] OR "interleukin-27"[All Fields] OR "interleukin 27"[All Fields] OR "interleukin-23"[MeSH Terms] OR "interleukin-23"[All Fields] OR "interleukin 23"[All Fields] |
| 5. | "tumour necrosis factor receptors"[All Fields] OR "receptors, tumor necrosis factor"[MeSH Terms] OR "tumor necrosis factor receptors"[All Fields] |
| 6. | TNF antagonis*[Title/Abstract] OR TNF inhibit*[Title/Abstract] |
| 7. | T cell helper[Title/Abstract] |
| 8. | anti-TNF[Title/Abstract] |
| 9. | 1 OR 2 OR 3 OR 4 OR 5 OR 6 OR 7 OR 8 |

**Cochrane search terms**

| | |
|---|---|
| 1. | MeSH descriptor: [Antibodies, Monoclonal] explode all trees |
| 2. | MeSH descriptor: [Interleukin-12] explode all trees |
| 3. | MeSH descriptor: [Interleukin-17] explode all trees |
| 4. | MeSH descriptor: [Interleukin-23] explode all trees |
| 5. | MeSH descriptor: [Receptors, Tumor Necrosis Factor] explode all trees |
| 6. | adalimumab OR etanercept OR infliximab OR ixekizumab OR ustekinumab OR secukinumab:ti,ab,kw |
| 7. | enbrel or remicade or humira or stelara or cosentyx OR taltz:ti,ab |
| 8. | biosimilar OR biosimilars:ti,ab |
| 9. | MeSH descriptor:[biosimilar pharmaceuticals] explode all trees |
| 10. | (TNF near/1 (antagonis* or inhibit*)):ti,ab |
| 11. | anti-TNF:ti,ab |
| 12. | (#1 OR #2 OR #3 OR #4 OR #5 OR #6 OR #7 OR #8 OR #9 OR #10 OR #11) |

**In people with psoriasis (all types), what is the risk of serious infection of biologics (brodalumab, guselkumab, tildrakizumab, risankizumab, certolizumab pegol) compared with each other, with adalimumab, etanercept, infliximab, ixekizumab, secukinumab, ustekinumab, methotrexate or with placebo?**

Search constructed by combining the columns in the following table using the **and** Boolean operator

| Population | Intervention | Comparison | Study filter used | Date parameters |
|---|---|---|---|---|
| Psoriasis | Biologic therapy (new biologics) NOT Biologic therapy (original biologics) | | RCTs, SRs and Observational studies [Medline and EMBASE only] | All years – 07/09/2018 |

The original search results from the 2016 guidelines were also resifted for all papers relating to brodalumab, guselkumab, tildrakizumab, risankizumab and certolizumab pegol.

#### S1.4.2 Biologic therapy (new biologics)

**Medline and EMBASE search terms**

| | |
|---|---|
| 1. | EMB.EXACT("brodalumab") OR EMB.EXACT("guselkumab") OR EMB.EXACT("tildrakizumab") OR EMB.EXACT("risankizumab") OR EMB.EXACT("certolizumab pegol") OR AB,TI,IF(brodalumab OR guselkumab OR tildrakizumab OR risankizumab OR certolizumab PRE/0 pegol) |
| 2. | AB,TI(kyntheum OR tremfya OR ilumya OR ABBV-066 OR cimzia) |
| 3. | 1 OR 2 |

**PubMed search terms**

| | |
|---|---|
| 1. | brodalumab OR guselkumab OR tildrakizumab OR risankizumab OR certolizumab pegol OR kyntheum OR tremfya OR ilumya OR ABBV-066 OR cimzia |

**Cochrane search terms**

| | |
|---|---|
| 1. | brodalumab, OR guselkumab OR tildrakizumab OR risankizumab OR certolizumab pegol:ti,ab,kw |
| 2. | kyntheum OR tremfya OR ilumya OR ABBV-066 OR cimzia:ti,ab |
| 3. | #1 OR #2 |

**Table S2. Characteristics of included studies**

| Study | Number of study sites | RCT phase (wks.) | Interventions during RCT phase | Number of participants in treatment arm | Number of participants in placebo arm | Mean age ± SD yrs. | Proportion of participants with PsA (%) | Mean PASI ± SD | Prior exposure to biologic therapy | Exclusion of patients with history of serious infections | Outcome definition clearly stated | Outcome definition |
|---|---|---|---|---|---|---|---|---|---|---|---|---|
| **Trials primarily involving etanercept** | | | | | | | | | | | | |
| Bachelez *et al.* 2015<br><br>USA and Canada<br><br>Industry funded | 122 | 12 | ETA 50 mg SC. BIW | 336 | 108 | Median ETA 42; PBO 46 | ETA 21.2; PBO 21.2 | Median ETA 19.4; PBO 19.5 | ETA 11%, placebo 11% | Yes (within 6 mos.) | Yes | Serious infection |
| Bagel *et al.* 2012<br><br>USA<br><br>Industry funded | Multicentre, NR | 12 | ETA 50 mg SC. BIW | 62 | 62 | Median ETA 39; PBO 42 | NR | Median ETA 15.2; PBO 15.5 | ETA 6.8% (anti-TNF), 3.2% (non-anti-TNF), placebo 6.5% (anti-TNF), 4.8% (non-anti-TNF) | NR | No | Individual serious adverse event |
| Gottlieb *et al.* 2011<br><br>USA<br><br>Industry funded | 33 | 12 | ETA 50 mg SC. BIW | 141 | 68 | ETA 43.1±12.5; PBO 44±13.6 | ETA 22.7; PBO 20.6 | ETA 19.4±8; PBO 18.5±6.9 | ETA 14.2%, placebo 14.7% | NR | No | Serious infection |
| Paller *et al.* 2008<br><br>USA<br><br>Industry funded | 42 | 12 | ETA 0.8 mg/kg | 106 | 105 | Median ETA 14, PBO 13 | ETA 5; PBO 14 | ETA 16.7; PBO 16.4 | No | NR | No | Serious infection |
| Papp *et al.* 2005<br><br>Canada and Western Europe<br><br>Industry funded | 50 | 12 | ETA 25 mg SC. BIW; ETA 50 mg SC. BIW | 196, 194 | 193 | 25 mg 46; 50 mg 44.5; PBO 44 | 25 mg 54%; 50 mg 50%; PBO 50% | 25 mg 16.9; 50 mg 16.1; PBO 16 | No | Yes (within 1 mo.) | No | Serious infectious event |

| Study | Number of study sites | RCT phase (wks.) | Interventions during RCT phase | Number of participants in treatment arm | Number of participants in placebo arm | Mean age ± SD yrs. | Proportion of participants with PsA (%) | Mean PASI ± SD | Prior exposure to biologic therapy | Exclusion of patients with history of serious infections | Outcome definition clearly stated | Outcome definition |
|---|---|---|---|---|---|---|---|---|---|---|---|---|
| Reich JEADV 2017 LIBERATE<br><br>Worldwide<br><br>Industry funded | Multicentre, NR | 16 | ETA 50 mg QW | 83 | 84 | ETA 50 mg; 47.0±14.1 | NR | ETA 50 mg 20.3±7.9; PBO 19.4±6.8 | No | Yes | Yes | Serious opportunistic infections |
| Strober et al 2011<br><br>USA<br><br>Industry funded | 41 | 12 | ETA 50 mg SC. BIW | 139 | 72 | ETA 45.2±14.8; PBO 45.0±13.9 | ETA 33.1; PBO 20.8 | ETA 18.5±6.0; PBO 18.3±6.4 | ETA 7.9%, placebo 4.2% | NR | No | Serious infection |
| Tyring *et al.* 2006<br><br>USA and Canada<br><br>Industry funded | 39 | 12 | ETA 50 mg SC. BIW | 311 | 309 | ETA 45.8 ±12.8; PBO 45.6±12.1 | ETA 35; PBO 32.6 | ETA 18.3±7.6; PBO 18.1±7.4 | No | NR | No | Serious infection |
| Van de Kerkhof *et al.* 2008<br><br>Belgium, France, Germany, Hungary, Italy, the Netherlands, Poland, Romania, and Spain<br><br>Industry funded | NR | 12 | ETA 50 mg SC. QW | 96 | 46 | ETA 45.9 ±12.8; PBO 43.6±12.6 | ETA 15.6; PBO 10.9 | ETA 21.4±9.3; PBO 21.0±8.7 | NR | Yes (within 1 mo.) | No | Serious infection |
| **Trials primarily involving infliximab** | | | | | | | | | | | | |
| Yang *et al.* 2012<br><br>China<br><br>Funding, NR | 9 | 10 | INF 5 mg/kg IV drip infusion wk. 0, 2 and 6 | 84 | 45 | INF 39.4±12.3; PBO 40.1±11.1 | NR | INF 23.9±10.7; PBO 25.3±12.7 | NR | Yes (within 2 mos.) | No | Individual serious adverse event |

| Study | Number of study sites | RCT phase (wks.) | Interventions during RCT phase | Number of participants in treatment arm | Number of participants in placebo arm | Mean age ± SD yrs. | Proportion of participants with PsA (%) | Mean PASI ± SD | Prior exposure to biologic therapy | Exclusion of patients with history of serious infections | Outcome definition clearly stated | Outcome definition |
|---|---|---|---|---|---|---|---|---|---|---|---|---|
| **Trials primarily involving adalimumab** | | | | | | | | | | | | |
| Asahina *et al.* 2010<br><br>Japan<br><br>Industry funded | 42 | 16 (SAEs reported at wk. 24) | ADA 40 mg eow; ADA 80 mg at wk. 0 and then 40 mg eow; 80 mg eow | 40 mg eow 38; 40 mg eow + 80 mg loading dose 43; 80 mg eow 42 | 46 | 40 mg eow 47.8±12.81; 40 mg eow + 80 mg loading dose 44.2±14.32; 80 mg eow 43.5±12.4; PBO 43.9±10.75 | NR | 40 mg eow 25.44±8.9 77; 40 mg eow + 80 mg loading dose 30.24±10. 946; 80 mg eow 28.27±11. 029; PBO 29.10±11. 767 | NR | NR | No | Individual serious adverse event |
| Cai *et al.* 2016<br><br>China<br><br>Industry funded | 16 | 12 | ADA 40 mg EOW | 338 | 87 | ADA 43.1±11.91; PBO 43.8±12.45 | ADA 12.7; PBO 11.5 | ADA 28.2 ± 12.0; PBO 25.6 ± 10.98 | No | Yes (within 1 mo.) | Yes | Serious infection |
| Gordon *et al.* 2006<br><br>USA and Canada<br><br>Industry funded | 18 | 12 | ADA 40 mg EOW; ADA 40 mg weekly | EOW 46; weekly 50 | 52 | 44 | EOW 31; weekly 33; PBO 24 | EOW 16; weekly 16.7; PBO 14.5 | No | NR | No | Infectious serious adverse event |
| Papp et al, Lancet 2017<br><br>Belgium, Canada, Czech Republic, Germany, Hungary, Italy, Mexico, Netherlands, Poland, Spain, Switzerland, and Turkey | 38 | 16 | ADA 0.8 mg/kg; ADA 0.4 mg/kg; MTX | ADA 0.8 mg/kg 38; ADA 0.4 mg/kg 39; MTX 37 | NA | ADA 0.8 mg/kg 13.0 (3.3); ADA 0.4 mg/kg 12.6 (4.4); MTX 13.4 (3.5) | NR | ADA 0.8 mg/kg 18.9 (10.0); ADA 0.4 mg/kg 16.9 (5.8); MTX 19.2 (10.0) | ADA 0.8 mg/kg 10.0%; ADA 0.4 mg/kg 5.8%; MTX 10.0% | Yes | Yes (within 30 days for IV treatment or within 14 days for oral treatment) | Serious infection |

| Study | Number of study sites | RCT phase (wks.) | Interventions during RCT phase | Number of participants in treatment arm | Number of participants in placebo arm | Mean age ± SD yrs. | Proportion of participants with PsA (%) | Mean PASI ± SD | Prior exposure to biologic therapy | Exclusion of patients with history of serious infections | Outcome definition clearly stated | Outcome definition |
|---|---|---|---|---|---|---|---|---|---|---|---|---|
| Industry funded | | | | | | | | | | | | |
| Menter *et al.* 2008<br><br>USA and Canada<br><br>Industry funded | 81 | 16 | ADA 80 mg SC. at wk. 0 followed by 40 mg SC. EOW starting at wk.1 | 814 | 398 | ADA 44.1±13.2; PBO 45.4±13.4 | ADA 27.5; PBO 28.4 | ADA 19.0±7.08; PBO 18.8±7.09 | ADA 11.9%, placebo 13.3% | NR | No | Serious infectious adverse event |
| Saurat *et al.* 2008<br><br>Canada and Europe<br><br>Industry funded | 28 | 16 | ADA 80 mg SC. at wk. 0 followed by 40 mg SC. EOW starting at wk. 1; MTX 7.5 mg orally increased as needed and tolerated to 25 mg weekly | ADA 108; MTX 110 | PBO 53 | ADA 42.9±12.6; MTX 41.6±12.0; PBO 40.7±11.4 | ADA 21.3; MTX 17.3; PBO 20.8 | ADA 20.2±7.5; MTX 19.4±7.4; PBO 19.2±6.9 | No | NR | No | Serious infection |
| **Trials primarily involving ustekinumab** | | | | | | | | | | | | |
| Griffiths *et al.* 2010<br><br>Worldwide<br><br>Industry funded | 67 | 12 | UST SC. 45 mg or 90 mg at wk. 0 and 4; ETA 25 mg SC. BIW | UST 45 mg 209; UST 90 mg 347; ETA 347 | NA | UST 45 mg 45.1±12.6; 90 mg 44.8±12.3; ETA 45.7±13.4 | UST 45 mg 29.7; 90 mg 27.4; ETA 27.4 | UST 45 mg 20.5±9.2; 90 mg 19.9±8.4; ETA 18.6±6.2 | (INF, ADA, alefacept, efalizumab): ETA 11.8%, UST 45 mg 12.4%, UST 90 mg 10.4% | Yes | No | Infections under serious adverse events |

| **Study** | **Number of study sites** | **RCT phase (wks.)** | **Interventions during RCT phase** | **Number of participants in treatment arm** | **Number of participants in placebo arm** | **Mean age ± SD yrs.** | **Proportion of participants with PsA (%)** | **Mean PASI ± SD** | **Prior exposure to biologic therapy** | **Exclusion of patients with history of serious infections** | **Outcome definition clearly stated** | **Outcome definition** |
|---|---|---|---|---|---|---|---|---|---|---|---|---|
| Igarashi *et al.* 2012<br><br>Japan<br><br>Industry funded | 35 | 12 | UST SC. 45 mg or 90 mg at wk. 0 and 4 | 45 mg 64; 90 mg 62 | 32 | UST 45 mg 46.6±12.5; UST 90 mg 46.8±12.8; PBO 48.5±12.7 | 45 mg 9.4; 90 mg 11.3; PBO 3.1 | 45 mg 30.1±12.9; 90 mg 28.7±11.2; PBO 30.3±11.8 | UST 45 mg 1.6%, UST 90 mg 0%, placebo 0% | Yes | No | Serious infection |
| Landells *et al.* 2015<br><br>Canada and Europe<br><br>Industry funded | 36 | 12 | UST - standard dose 0.75 mg/kg for weight </= 60kg; same as adult dose for weight > 60kg; half dose 0.375 mg/kg </= 60kg; 22.5 mg >60kg to < 100kg; 45 mg for > 100kg | 73 | 37 | UST 14.9±1.7 PBO 15.6±1.5 | NR | UST 21.3±9.4 PBO 20.8±8.0 | UST 8.3%, UST 10.8%, placebo 13.5% | NR | No | Serious infection |
| Leonardi *et al.* 2008<br><br>USA, Canada, Belgium<br><br>Industry funded | 48 | 12 | UST SC. 45 mg or 90 mg at wk. 0 and 4 | UST 45 mg 255; UST 90 mg 256 | 255 | 45 mg 44.8±12.5; 90 mg 46.2±11.3 | 45 mg 29; 90 mg 37.1; PBO 35.3 | 45 mg 20.5±8.6; 90 mg 19.7±7.6; PBO 20.4±8.6 | (ETA , alefacept, efalizumab, INF, ADA): UST 45 mg 52·5%, UST 90 mg 50·8%, placebo 50·2% | Yes | No | Serious infection |
| Papp *et al.* 2008 | 70 | 12 | UST SC. 45 mg or 90 mg | 45 mg 409; 90 mg 411 | 410 | 45 mg 45.1±12.1; | 45 mg 26.2; 90 mg 22.9; | 45 mg 19.4±6.8; | (ETA , alefacept, | Yes | No | Serious infection |

| Study | Number of study sites | RCT phase (wks.) | Interventions during RCT phase | Number of participants in treatment arm | Number of participants in placebo arm | Mean age ± SD yrs. | Proportion of participants with PsA (%) | Mean PASI ± SD | Prior exposure to biologic therapy | Exclusion of patients with history of serious infections | Outcome definition clearly stated | Outcome definition |
|---|---|---|---|---|---|---|---|---|---|---|---|---|
| Europe and USA<br><br>Industry funded | | | at wk. 0 and 4 | | | 90 mg 46.26±12.1 ; PBO 47.0±12.5 | PBO 25.6 | 90 mg 20.1±7.5; PBO 19.4±7.5 | efalizumab, INF, ADA): UST 45 mg 38.4%, UST 90 mg 36.5%, placebo 38.8% | | | |
| Tsai *et al.* 2011<br><br>Korea and Taiwan<br><br>Industry funded | 13 | 12 | UST SC. 45 mg at wk. 0 and 4 | 61 | 60 | UST 40.9±12.7; PBO 40.4±10.1 | UST 16.4; PBO 11.7 | UST 25.2±11.9; PBO 22.9±8.6 | (ETA , efalizumab, INF, ADA): UST 21.3%; placebo 15.0% | NR | No | Serious infection |
| Zhu *et al.* 2013<br><br>China<br><br>Industry funded | 14 | 12 | UST SC. 45 mg at wk. 0 and 4 | 160 | 162 | UST 40.1±12.4; PBO 39.2±12.2 | UST 8.8; PBO 8.6 | UST 23.2±9.5; PBO 22.7±9.5 | UST 11.9%, placebo 6.8% | NR | No | Serious infection |
| **Trials primarily involving secukinumab** | | | | | | | | | | | | |
| Blauvelt *et al.* 2015<br><br>North America and Europe<br><br>Industry funded | 32 | 12 | SEC 150 mg or 300 mg SC. at wk. 0, 1, 2, 3, 4 and then every 4 wks. | 150 mg 59; 300 mg 59 | 59 | 150 mg 46.0±15.09; 300 mg 45.1±12.57; PBO 46.5±14.14 | NR | 150 mg 21.4±9.1; 300 mg 20.7±7.95; PBO 21.1±8.49 | SEC 300 mg 39.0% (39.1% failed), SEC 150 mg 47.5% (64.3% failed), placebo 44.1% (53.8% failed) | Yes (within 2 wks.) | No | Serious infection |
| Langley et 2014 (FIXTURE)<br><br>Worldwide<br><br>Industry funded | 231 | 12 | SEC 150 mg or 300 mg SC. at wk. 0, 1, 2, 3, 4 and then every 4 | SEC 150 mg 327; SEC 300 mg 327; ETA 323 | 326 | SEC 150 mg 45.4±12.9; SEC 300 mg 44.5±13.2; ETA 43.8±13.0; | SEC 150 mg 15.0; SEC 300 mg 15.3; ETA 13.5; PBO 15.0 | SEC 150 mg 23.7±10.5; SEC 300 mg 23.9±9.9; ETA | SEC 300 mg 11.6%, SEC 150 mg 13.8%, ETA 13.8%, | NR | No | Serious infection* |

| Study | Number of study sites | RCT phase (wks.) | Interventions during RCT phase | Number of participants in treatment arm | Number of participants in placebo arm | Mean age ± SD yrs. | Proportion of participants with PsA (%) | Mean PASI ± SD | Prior exposure to biologic therapy | Exclusion of patients with history of serious infections | Outcome definition clearly stated | Outcome definition |
|---|---|---|---|---|---|---|---|---|---|---|---|---|
| | | | wks.; ETA 50 mg SC BIW | | | PBO 44.1±12.6 | | 23.2±9.8; PBO 24.1±10.5 | placebo 10.7% | | | |
| Langley *et al.* 2014 (ERASURE)<br><br>Worldwide<br><br>Industry funded | 88 | 12 | SEC 150 mg or 300 mg SC. at wk. 0, 1, 2, 3, 4 and then every 4 wks. | 150 mg 245; 300 mg 245 | 248 | 150 mg 44.9±13.3; 300 mg 44.9±13.5; PBO 45.4±12.6 | 150 mg 18.8; 300 mg 23.3; PBO 27.4 | 150 mg 22.3±9.8; 300 mg 22.5±9.2; PBO 21.4±9.1 | SEC 300 mg 28.6%, SEC 150 mg 29.8%, placebo 29.4% | NR | No | Serious infection* |
| Paul *et al.* 2015<br><br>Worldwide<br><br>Industry funded | 38 | 12 | SEC 150 mg or 300 mg SC. at wk. 0, 1, 2, 3, 4 and then every 4 wks. | 150 mg 61; 300 mg 60 | 61 | 150 mg 43.9±14.41; 300 mg 46.6±14.23; PBO 43.7±12.74 | 150 mg 26.2; 300 mg 23.3; PBO 19.7 | 150 mg 22.0±8.85; 300 mg 18.9±6.37; PBO 19.4±6.70 | SEC 300 mg 25.0%, SEC 150 mg 24.6%, placebo 21.3% | NR | No | Serious infection |
| Rich *et al.* 2013<br><br>France, Germany, Iceland, Israel, Japan, Norway, USA<br><br>Industry funded | 60 | 12 | SEC x1 150 mg SC; 150 mg SC at 0, 4, 8 wk. (monthly); 150 mg SC at 0, 1, 2, 4 Wks. (early) | 150 mg x1 66; 150 mg monthly 138; 150 mg early 133 | 67 | 150 mg x1 42.7; 150 mg monthly 44.2; 150 mg early 44.5; PBO 44.2 | 150 mg x1 22.7; 150 mg monthly 32.6; 150 mg early 29.3; PBO 17.9 | 150 mg x1 19.9±6.73; 150 mg monthly 20.8±8.08; 150 mg early 19.9±7.81; PBO 20.5±9.31 | SEC single 31.8%, SEC monthly 29.7%, SEC early 30.1%, placebo 25.4% | NR | No | Serious infection |
| Thaci *et al.* 2015<br><br>Worldwide<br><br>Industry funded | Multicentre | 16 | SEC 300 mg SC. at wk. 0, 1, 2, 3, 4 and then every 4 wks.; UST SC. 45 mg or 90 mg at wk. 0 and 4 | SEC 337; UST 339 | NA | SEC 45.2±13.96; UST 44.6±13.67 | SEC 20.5; UST 15.9 | SEC 21.7±8.5; UST 21.5±8.07 | UST 13.0% (10.0% failed), SEC 14.2% (10.7% failed) | NR | No | Serious infection* |
| **Studies primarily involving ixekizumab** | | | | | | | | | | | | |

| Study | Number of study sites | RCT phase (wks.) | Interventions during RCT phase | Number of participants in treatment arm | Number of participants in placebo arm | Mean age ± SD yrs. | Proportion of participants with PsA (%) | Mean PASI ± SD | Prior exposure to biologic therapy | Exclusion of patients with history of serious infections | Outcome definition clearly stated | Outcome definition |
|---|---|---|---|---|---|---|---|---|---|---|---|---|
| Gordon *et al.* 2016<br><br>Worldwide<br><br>Industry funded | 100 | 12 | U1<br>IXE 160 mg at wk. 0 then 80 mg every 4 wks.; IXE 160 mg at wk. 0 then 80 mg every 2 wks.<br><br>U2 and U3<br>IXE 160 mg at wk. 0 then 80 mg every 4 wks.; IXE 160 mg at wk. 0 then 80 mg every 2 wks.; ETA 50 mg twice a wk. | IXE every 4 wks. U1 432; U2 347; U3 386<br><br>IXE every 2 wks.: U1 433; U2 351; U3 385<br><br>ETA U2 358; U3 382 | U1 431; U2 168; U3 193 | IXE every 4 wks.: U1 46±13; U2 45±14; U3 46±13<br><br>IXE every 2 wks.: U1 45±12; U2 45±13; U3 46±13<br>ETA U2 45±13; U3 46±14<br>PBO U2 45±12; U3 46±12 | NR | IXE every 4 wks.: U1 20±7; U2 20±7; U3 21±8<br>IXE every 2 wks.: U1 20±8; U2 19±7; U3 21±8<br>ETA U2 19±7; U3 21±8<br>PBO U1 20±9; U2 21±8; U3 21±8 | IXE every 4 weeks 38.9%, IXE every 2 weeks 40.0%, placebo 42.0% (U1), IXE every 4 weeks 25%, IXE every 2 weeks 24%, ETA 21%, placebo 26% (U2), IXE every 4 weeks 15%, IXE every 2 weeks 15%, ETA 16%, placebo 17% (U3) | NR | Yes | Serious infection |
| Griffiths *et al.* 2015<br><br>Worldwide<br><br>Industry funded | 126 | 12 | IXE 160 mg at wk. 0 then 80 mg every 4 wks.; IXE 160 mg at | IXE every 4 wks. U2 347; U3 386<br><br>IXE every 2 wks.: U2 | U2 168; U3 193 | IXE every 4 wks.:<br>U2 45±14;<br>U3 46±13<br><br>IXE every 2 | NR | IXE every 4 wks.<br>U2 20±7;<br>U3 21±8<br><br>IXE every 2 | IXE every 4 weeks 25%, IXE every 2 weeks 24%, ETA 21%, placebo 26% | NR | No | Serious infection* |

| Study | Number of study sites | RCT phase (wks.) | Interventions during RCT phase | Number of participants in treatment arm | Number of participants in placebo arm | Mean age ± SD yrs. | Proportion of participants with PsA (%) | Mean PASI ± SD | Prior exposure to biologic therapy | Exclusion of patients with history of serious infections | Outcome definition clearly stated | Outcome definition |
|---|---|---|---|---|---|---|---|---|---|---|---|---|
| | | | wk. 0 then 80 mg every 2 wks. ETA 50 mg SC. BIW | 351; U3 385<br><br>ETA U2 358; U3 382 | | wks.: U2 45±13; U3 46±13<br><br>ETA U2 45±13; U3 46±14<br><br>PBO U2 45±12; U3 46±12 | | wks. U2 19±7; U3 21±8<br><br>ETA U2 19±7; U3 21±8<br><br>PBO U2 21±8; U3 21±8 | (U2), IXE every 4 weeks 15%, IXE every 2 weeks 15%, ETA 16%, placebo 17% (U3) | | | |
| Leonardi *et al.* 2012<br><br>USA and Denmark<br><br>Industry funded | Multicentre | 12 | IXE 10 mg/IXE 25 mg/IXE 75 mg/IXE 150 mg at wk. 0, 2, 4 and every 4 wks. | IXE 10 mg 28;<br>IXE 25 mg 30;<br>IXE 75 mg 29;<br>IXE 150 mg 28 | 27 | IXE 10 mg 48±11<br>IXE 25 mg 46±15<br>IXE 75 mg 46±13<br>IXE 150 mg 46±13<br>PBO 45±13 | NR | IXE 10 mg 19.2±8.0;<br>IXE 25 mg 18.6±4.9;<br>IXE 75 mg 17.2±4.3;<br>IXE 150 mg 17.7±6.2;<br>PBO 16.5±5.3 | NR | Yes | Yes | Serious infection |
| **Studies primarily involving brodalumab** | | | | | | | | | | | | |
| Lebwohl *et al.* 2015<br><br>Worldwide<br><br>Industry funded | 142 | 12 | UST 45 mg for patients with a body weight ≤100 kg and 90 mg for patients with a body | UST AMAGINE-2 300; AMAGINE-3 313<br><br>BRO 140 mg AMAGINE-2 610; AMAGINE-3 629 | AMAGINE-2, 309; AMAGINE-3, 315 | AMAGINE-2 45±13; AMAGINE-3 45±13 | AMAGINE-2 19; AMAGINE-3 18 | AMAGINE-2 20.3±8.2; AMAGINE-3 20.2±8.4 | AMAGINE – 2, 29%; AMAGINE – 3, 25% | NR | Yes | Serious infection |

| Study | Number of study sites | RCT phase (wks.) | Interventions during RCT phase | Number of participants in treatment arm | Number of participants in placebo arm | Mean age ± SD yrs. | Proportion of participants with PsA (%) | Mean PASI ± SD | Prior exposure to biologic therapy | Exclusion of patients with history of serious infections | Outcome definition clearly stated | Outcome definition |
|---|---|---|---|---|---|---|---|---|---|---|---|---|
| | | | weight >100 kg at wk. 0, 4 and every 12 wks.; BRO 140 mg at wk. 0, 1, 2 and every 2 wks.; BRO 210 mg at wk. 0, 1, 2 and every 2 wks. | BRO 210 mg AMAGINE-2 612; AMAGINE-3 624 | | | | | | | | |
| Papp *et al.* BJD 2016<br><br>Europe, Canada, and USA<br><br>Industry funded | 73 | 12 | BRO 140 mg; BRO 210 mg | BRO 140 mg, 219; BRO 210 mg, 222 | 220 | BRO 140 mg 46±13; BRO 210 mg 46±12; PBO 47±13 | BRO 140 mg 27; BRO 210 mg 26; PBO 29 | BRO 140 mg 20.0±7.4; BRO 210 mg 19.4±6.6 PBO; 19.7±7.7 | BRO 140 mg 45% BRO 210 mg 47% placebo 46% | Yes (within 8 wks.) | Yes | Serious infection |
| **Studies primarily involving guselkumab** | | | | | | | | | | | | |
| Blauvelt *et al.* 2017<br><br>Worldwide<br><br>Industry funded | 101 | 16 | GUS 100 mg at wk. 0 and 4, then every 8 wks.; ADA 80 mg at wk. 0 and then 40 mg every other | GUS 329; ADA 334 | 174 | GUS 43.9±12.74 ; ADA 42.9±12.58 | GUS 19.5; ADA 18.6; PBO 17.2 | GUS 22.1±9.49; ADA 22.4±8.97; PBO 20.4±8.74 | NR | NR | Yes | Serious infection |

| Study | Number of study sites | RCT phase (wks.) | Interventions during RCT phase | Number of participants in treatment arm | Number of participants in placebo arm | Mean age ± SD yrs. | Proportion of participants with PsA (%) | Mean PASI ± SD | Prior exposure to biologic therapy | Exclusion of patients with history of serious infections | Outcome definition clearly stated | Outcome definition |
|---|---|---|---|---|---|---|---|---|---|---|---|---|
| | | | wk. starting at wk. 1 | | | | | | | | | |
| Gordon *et al.* 2015<br><br>USA and Europe<br><br>Industry funded | 43 | 16 | GUS 5 mg at wk. 0 and 4 then every 12 wks.; 50 mg at wk. 0 and 4 then every 12 wks.; 100 mg at wk. 0 and 4 then every 12 wks.; 200 mg at wk. 0 and 4 then every 12 wks.<br><br>ADA 80 mg at wk. 0 and then 40 mg every other wk. starting at wk.1 | GUS 208; ADA 43 | 42 | Median GUS 44.0; ADA 50.0; PBO 46.5 | GUS 25; ADA 25.6; PBO 28.6 | ADA 20.2±7.58; PBO 21.8±9.98 | ADA 60%, placebo 26% | Yes (within 2 mos.) | No | Serious infection |
| Reich *et al.* JAAD 2017<br><br>Worldwide<br><br>Industry funded | 115 | 16 | GUS 100 mg at wk. 0 and 4 then every 8 wks.; ADA 80 mg at wk. 0 and then 40 mg every other wk. starting at wk. 1 | GUS 496; ADA 248 | 248 | GUS 43.7±12.2; ADA 43.2±11.9 | GUS 17.9; ADA 17.7; PBO 18.5 | GUS 21.9 ± 8.8; ADA 21.7 ± 9.0; PBO 21.5 ± 8.0 | NR | Yes | No | Serious infection |

| Study | Number of study sites | RCT phase (wks.) | Interventions during RCT phase | Number of participants in treatment arm | Number of participants in placebo arm | Mean age ± SD yrs. | Proportion of participants with PsA (%) | Mean PASI ± SD | Prior exposure to biologic therapy | Exclusion of patients with history of serious infections | Outcome definition clearly stated | Outcome definition |
|---|---|---|---|---|---|---|---|---|---|---|---|---|
| **Studies primarily involving certolizumab pegol** | | | | | | | | | | | | |
| Gottlieb *et al.* JAAD 2018 CIMPASI 1<br><br>North America and Europe<br><br>Industry funded | Multicentre, NR | 16 | CZP 200 mg every 2 wks.; CZP 400 mg every 2 wks. | CZP 200 mg 95; CZP 400 mg 88 | 51 | CZP 200 mg 44.5±13.1; CZP 400 mg 43.6±12.1; PBO 47.9±12.8 | CZP 200 mg 10.5; CZP 400 mg 17.0; PBO 7.8 | CZP 200 mg 20.1±8.2; CZP 400 mg 19.6±7.9; PBO 19.8±7.5 | No | Yes | Yes | Serious infection |
| Gottlieb *et al.* 2018 JAAD CIMPASI 2<br><br>North America and Europe<br><br>Industry funded | Multicentre, NR | 16 | CZP 200 mg every 2 wks.; CZP 400 mg every 2 wks. | CZP 200 mg every 2 wks. 91; CZP 400 mg every 2 wks. 87 | 49 | CZP 200 mg 45.6±13.2; CZP 400 mg 45.0±12.9; PBO 45.7 ±13.8 | CZP 200 mg 24.2; CZP 400 mg 29.9; PBO 18.4 | CZP 200 mg 18.4±5.9; CZP 400 mg 19.5±6.7; PBO 17.3±5.3 | No | Yes | Yes | Serious infection |
| Lebwohl *et al.* 2018<br><br>North America and Europe<br><br>Industry funded | 559 | 12 | CZP 200 mg every 2 wks.; CZP 400 mg every 2 wks.; ETA 50 mg BIW | CZP 200 mg every 2 wks. 165; CZP 400 mg every 2 wks. 167; ETA 170 | 57 | CZP 200 mg 46.7±13.5; CZP 400 mg 45.4±12.4; ETA 44.6±14.1; PBO 46.5±12.5 | CZP 200 mg every 2 wks. 16.4; CZP 400 mg every 2 wks. 14.4; ETA 15.9; PBO 21.1 | CZP 200 mg every 2 wks. 21.4±8.8; CZP 400 mg every 2 wks. 20.8±7.7; ETA 21.0±8.2; PBO 19.1±7.1 | NR | Yes | Yes | Serious infections and infestations |
| Reich *et al.* 2012<br><br>France and Germany<br><br>Industry funded | 15 | 12 | CZP 400 mg at wk. 0 then every 2 wks. until wk. 10; CZP 400 mg at wk. 0 then 200 mg every 2 wks. | CZP 200 mg 59; CZP 400 mg 58 | 59 | CZP 400 mg 43.6±12.4; CZP 200 mg 43.3±10.1; PBO 43.3±12.8 | NR | CZP 400 mg 22.0 ±8.1; CZP 200 mg 21.4±8.2; PBO 22.6±8.8 | No | Yes | No | Individual serious adverse event |

| Study | Number of study sites | RCT phase (wks.) | Interventions during RCT phase | Number of participants in treatment arm | Number of participants in placebo arm | Mean age ± SD yrs. | Proportion of participants with PsA (%) | Mean PASI ± SD | Prior exposure to biologic therapy | Exclusion of patients with history of serious infections | Outcome definition clearly stated | Outcome definition |
|---|---|---|---|---|---|---|---|---|---|---|---|---|
| | | | until wk. 10 | | | | | | | | | |
| **Studies primarily involving tildrakizumab** | | | | | | | | | | | | |
| Papp *et al.* 2015<br><br>USA, Canada, Japan, and Europe<br><br>Industry funded | 64 | 12 | TIL 5 mg at wk. 0 and 4, then every 12 wks.; TIL 25 mg at wk. 0 and 4, then every 12 wks.; TIL 100 mg at wk. 0 and 4, then every 12 wks.; TIL 200 mg at wk. 0 and 4, then every 12 wks. | TIL 5 mg 42; TIL 25 mg 92; TIL 100 mg 89; TIL 200 mg 86 | 46 | TIL 5 mg 43.2±12.9; TIL 25 mg 46.3±13.7; TIL 100 mg 45.5±12.8; TIL 200 mg 43.2±12.6 | TIL 5 mg 19; TIL 25 mg 16; TIL 100 mg 17; TIL 200 mg 17 | NR | TIL 5 mg 26% TIL 25 mg 28% TIL 100 mg 26% TILL 200 mg 26% placebo 28% | Yes | Yes | Serious infection |
| Reich *et al.* 2017 reSURFACE1<br><br>Australia, Canada, Japan, UK, and the USA<br><br>Industry funded | 118 | 12 | TIL 100 mg SC. wk. 0, then every 4 wks.; TIL 200 mg SC. wk. 0, then every 4 wks. | TIL 100 mg 309; TIL 200 mg 308 | 155 | TIL 100 mg 46.4 ± 13.1; TIL 200 mg 46.9 ± 13.2; PBO 47.9 ± 13.5 | NR | TIL 100 mg 20.0±7.85; TIL 200 mg 20.7±8.51; PBO 19.3±7.07 | TIL 200 mg 23% TIL 100 mg 23% placebo 23% | Yes (within 2 wks.) | Yes | Serious infection |
| Reich *et al.* 2017 reSURFACE 2 | 132 | 12 | TIL 100 mg wk. 0 then every 4 wks. TIL 200 mg | TIL 100 mg 307; TIL 200 mg 314; | 156 | TIL 100 mg 44.6±13.6; TIL 200 mg 44.6±13.6; | NR | TIL 100 mg 20.5±7.63; TIL 200 mg 19.8±7.52; | TIL 200 mg 12% TIL 100 mg 13% ETA | Yes (within 2 wks.) | Yes | Serious infection |

| Study | Number of study sites | RCT phase (wks.) | Interventions during RCT phase | Number of participants in treatment arm | Number of participants in placebo arm | Mean age ± SD yrs. | Proportion of participants with PsA (%) | Mean PASI ± SD | Prior exposure to biologic therapy | Exclusion of patients with history of serious infections | Outcome definition clearly stated | Outcome definition |
|---|---|---|---|---|---|---|---|---|---|---|---|---|
| Austria, Belgium, Canada, Czech Republic, Denmark, France, Germany, Hungary, Italy, Israel, Netherlands, Poland, and the USA<br><br>Industry funded | | | wk. 0 then every 4 wks.; ETA 50 mg BIW | ETA 313 | | ETA 45.8±14.0 | | ETA 20.2±7.36; PBO 20±7.57 | 12% placebo 13% | | | |
| **Studies primarily involving risankizumab** | | | | | | | | | | | | |
| Gordon *et al.* 2018 UltIMMa-1<br><br>Australia, Austria, Belgium, Canada, Czech Republic, France, Germany, Japan, Mexico, Poland, Portugal, South Korea, Spain, and the USA<br><br>Industry funded | 139 | 16 | RIS 150 mg at wk. 0 and 4 then every 12 wks.; UST 45 mg at wk. 0 and 4 then every 12 wks. | RIS 150 mg 304; UST 45 mg 100 | 102 | RIS 48.3±13.4; UST 46.5±13.4; PBO 49.3±13.6 | RIS 28; UST 23; PBO 35 | RIS 20.6±7.7; UST 20.1±6.8; PBO 20.5±6.7 | RIS 34% UST 30% placebo 39% | Yes (chronic or acute infection) | Yes | Serious infection |
| Gordon *et al.* 2018 UltIMMa-2<br><br>Australia, Austria, Belgium, Canada, Czech Republic, France, Germany, Japan, Mexico, Poland, Portugal, | 139 | 16 | RIS 150 mg at wk. 0 and 4 then every 12 wks.; UST 45 mg at wk. 0 and 4 then every 12 wks. | RIS 150 mg 294; UST 45 mg 99 | 98 | RIS 46.2±13.7; UST 48.6±14.8; PBO 46.3±13.3 | RIS 25; UST 27; PBO 33 | RIS 20.5±7.8; UST 18.2±5.9; PBO 18.9±7.3 | RIS 40%; UST 43%; placebo 43% | Yes (chronic or acute infection) | Yes | Serious infection |

| **Study** | **Number of study sites** | **RCT phase (wks.)** | **Interventions during RCT phase** | **Number of participants in treatment arm** | **Number of participants in placebo arm** | **Mean age ± SD yrs.** | **Proportion of participants with PsA (%)** | **Mean PASI ± SD** | **Prior exposure to biologic therapy** | **Exclusion of patients with history of serious infections** | **Outcome definition clearly stated** | **Outcome definition** |
|---|---|---|---|---|---|---|---|---|---|---|---|---|
| South Korea, Spain, and the USA<br><br>Industry funded | | | | | | | | | | | | |
| Papp *et al.* 2017<br><br>North America and Europe<br>Industry funded | 32 | 12 | RIS 18 mg<br>RIS 90 mg<br>RIS 180 mg<br>UST | RIS 18 mg 43;<br>RIS 90 mg 41;<br>RIS 180 mg 42;<br>UST 40 | NA | RIS 18 mg 44±14;<br>RIS 90 mg 49±13;<br>RIS 180 mg 45±14;<br>UST 45±12 | RIS 18 mg 7;<br>RIS 90 mg 13;<br>RIS 180 mg 12;<br>UST 14 | RIS 18 mg 19±7;<br>RIS 90 mg 19±7;<br>RIS 180 mg 20±8;<br>UST 20±6 | RIS 18 mg 28%; RIS 90 mg 27%; RIS 180 mg 29%; UST 22% | Yes (chronic or acute infection) | Yes | Serious infection and infestation |

**Abbreviations:** ADA – adalimumab; BIW – twice weekly; EOW –every other week; ETA – etanercept; INF – infliximab; IV – intravenous; mos. – months; NA, not applicable; NR – not reported; PASI – psoriasis area and severity index; PBO – placebo; PsA – psoriatic arthritis; RCT – randomised controlled trial; SC – subcutaneous; SD – standard deviation; U2 – UNCOVER-2; U3 – UNCOVER-3; UST – ustekinumab; wks., weeks.

*Definition provided directly from sponsoring pharmaceutical company; not given in the published report. † incident population; ˜prevalent population; ‡ not clear if infection was serious

**Table S3. Excluded studies with reasons for exclusion**

| Reference | Reason for exclusion |
|---|---|
| Adenubiova, E. *J Dermatolog Treat* 2018; **29**:579-82. | Inappropriate study design – no comparator, retrospective |
| Armstrong, A. W. *Am J Clin Dermatol* 2016; **17**:691-9. | Sub-analysis of Revicki JDT (2007) and Menter JAAD (2008) |
| Asahina, A. *J Dermatol* 2016; **43**:1257-66. | Inappropriate study design – no comparator |
| Attia, A. *Clin Drug Investig* 2017; **37**:439-51. | SR screened for additional papers, none identified |
| Belinchón, I. *J Eur Acad Dermatol Venereol* 2017; **31**:1700-8. | Time point out of scope (7 years) |
| Bilal, J. *J Dermatolog Treat* 2018; **29**:569-78. | SR screened for additional papers, none identified |
| Bissonnette, R. *Br J Dermatol* 2017; **177**:1033-42. | Inappropriate comparison: same biologic different treatment regimens, original study excluded Mrowietz JAAD (2015) excluded for same reason |
| Blauvelt, A. *J Am Acad Dermatol* 2017; **77**:372-4. | Letter, looking at speed of efficacy outside scope, Lebwohl, *N Engl J Med* (2015) |
| Blauvelt, A. *J Am Acad Dermatol* 2017; **77**:855-62. | Inappropriate study design – no comparator |
| Blauvelt, A. *Br J Dermatol* 2018; **179**:623-31. | Inappropriate study design – no appropriate comparator |
| Blauvelt, A. *Br J Dermatol* 2018. | Pooled data, all original studies already included |
| Carneiro, C. *Dermatol Ther* 2017; **30**. | Time point out of scope (4 years) |
| Chen, Y. *Immunotherapy* 2015; **7**:1023-37. | Systemic review all original articles that meet our protocol already included |
| Correr, C. J. *Cad Saude Publica* 2013; **29 Suppl 1**:S17-31. | Not systematic, limited capture of outcomes |
| Cui, L. *Int Immunopharmacol* 2018; **62**:46-58. Erratum doi: 10.1016/j.intimp.2018.08.039. | SR screened for additional papers, none identified |
| de Vries, A. C. *Br J Dermatol* 2017; **176**:624-33. | Less than 25 patients on the ETA arm |
| Egeberg, A. *Br J Dermatol* 2018; **178**:509-19. | Outcomes – serious infection not reported |
| Foley, P. *JAMA Dermatol* 2018; **154**:676-83. | No outcomes not already extracted from Blauvelt JAAD (2017) and Reich JAAD (2017) |
| Garcia-Doval, I. *J Am Acad Dermatol* 2017; **76**:299-308.e16. | Inappropriate study design – doesn't separate biologics - no extractable data |
| Georgakopoulos, J. R. *J Eur Acad Dermatol Venereol* 2018; **32**:e32-e4. | Inappropriate study design – no comparator, retrospective – letter |
| Gniadecki, R. *J Eur Acad Dermatol Venereol* 2018; **32**:1297-304. | Inappropriate study design – no comparator |
| Gottlieb, A. B. (2003). Arch Dermatol 139: 1627-1632; discussion 1632. | Time point out of scope (20-30 weeks) |
| Gottlieb, A. B. (2004). J Am Acad Dermatol 51: 534-542. | Time point out of scope (20-30 weeks) |
| Gottlieb, A. B. *J Drugs Dermatol* 2016; **15**:1226-34. | Results for the placebo arm not reported after 12 weeks |
| Gordon, K.B. Br J Dermatol 2014; 170: 705-715 | Outcomes – serious infection not reported |
| Gottlieb, A. *J Am Acad Dermatol* 2017; **76**:70-80. | No outcomes not already extracted from Gottlieb JAAD (2016) |
| Griffiths, C. E. M. *Br J Dermatol* 2017; **176**:928-38. | Inappropriate study design – no appropriate comparator |
| Griffiths, C. E. M. *J Drugs Dermatol* 2018; **17**:826-32. | Inappropriate study design – no comparator after 52 weeks |
| Hjalte, F. *Br J Dermatol* 2018; **178**:245-52. | Inappropriate study design – doesn't separate biologics - no extractable data |
| Iskandar, I. Y. K. *Br J Dermatol* 2017; **177**:1410-21. | Data not in extractable format |
| Iskandar, I. Y. K. *J Invest Dermatol* 2018; **138**:775-84. | Data not in extractable format |
| Kalb, R. E. (2015). JAMA Dermatol 151: 961-969. | Intervention – non-biologic |
| Krueger, G. G. (2007). N Engl J Med 356: 580-592. | Time point out of scope (20-30 weeks) |
| Lacour, J. P. *J Eur Acad Dermatol Venereol* 2017; **31**:847-56. | No outcomes not already extracted from Paul JEADV (2015). Inappropriate study design – no comparator at 1 year. |
| Langley, R. G. *Br J Dermatol* 2018; **178**:114-23. | Out of scope for this question: inadequate response. |
| Leonardi, C. L. *J Eur Acad Dermatol Venereol* 2017; **31**:1483-90. | Pooled data, original studies already included; DLQI not reported as change from baseline |
| Leonardi, C. *J Am Acad Dermatol* 2018. | Inappropriate study design – no comparator |
| Lunder, T. *Biologicals* 2018; **54**:44-9. | Time point out of scope (10 years) |
| Lv, J. *Mol Pain* 2018; **14**:1744806918762205. | Inappropriate study design – doesn't separate biologics – screened for additional papers, none identified |

| | |
|---|---|
| Marinas, J. E. *Australas J Dermatol* 2018; **59**:e11-e4. | Outcomes – serious infection not reported |
| Menter, A. *Dermatol Ther (Heidelb)* 2017; **7**:365-81. | Inappropriate study design – no comparator |
| Menter, A. *J Eur Acad Dermatol Venereol* 2017; **31**:1686-92. | Pooled data, original studies already included |
| Menting, S. P. *Br J Dermatol* 2014; **171**:875-83. | Outcomes – serious infection not reported |
| Nakagawa, H. J Dermatol Sci 2016; 81:44-52. | Outcomes – serious infection not reported |
| Nemoto, O. *Br J Dermatol* 2018; **178**:689-96. | Does not meet protocol, less than 25 patients on each arm |
| No, D. J. *J Dermatolog Treat* 2018; **29**:460-6. | SR screened for additional papers, one identified Menting (2014) |
| No, D. J. *J Dermatolog Treat* 2018; **29**:467-74. | SR screened for additional papers, none identified |
| Ohtsuki, M. *J Dermatol* 2018; **45**:1053-62. | Outcomes – serious infection not reported |
| Papp, K. A. N Engl J Med 2012; 366:1181-9. | Outcomes – serious infection not reported |
| Papp, K. *J Am Acad Dermatol* 2015; **72**:436-9 e1. | Outcomes not reported in a way that can be extracted |
| Papp, K. A. *Br J Dermatol* 2017; **177**:1537-51. | Pooled data, all original studies already included |
| Papp, K. *Br J Dermatol* 2017; **177**:1562-74. | Inappropriate study design – no appropriate comparator |
| Papp, K. *J Am Acad Dermatol* 2017; **76**:1093-102. | Inappropriate study design – no appropriate comparator |
| Papp, K. A. *Br J Dermatol* 2018; **178**:674-81. | Pooled data, all original studies already included insufficient data for response rates for NAPSI |
| Papp, K. A. *J Am Acad Dermatol* 2018; **79**:277-86.e10. | Out of scope for this update |
| Papp, K. A. *J Cutan Med Surg* 2018. | Inappropriate study design – no comparator |
| Puig, L. *J Am Acad Dermatol* 2018; **78**:741-8. | No outcomes not already extracted from Blauvelt JAAD (2017) or Thaci JAAD (2015) |
| Paul, C. J Am Acad Dermatol 2018. | Outcome – serious infection not reported |
| Ramaekers, B. L. T. *Pharmacoeconomics* 2018; **36**:917-27. | Review |
| Reich, K. Br J Dermatol 2017; 177:1014-23. | Outcomes – serious infection not reported |
| Reich, K. *Arch Dermatol Res* 2015; **307**:875-83 | Data for individual biologics only represented graphically |
| Reich, K. (2005). Lancet 366: 1367-1374. | Time point out of scope (20-30 weeks) |
| Sbidian, E. *Cochrane Database Syst Rev* 2017; **12**:Cd011535. | SR screened for additional papers, one identified Bagel (2012) |
| Shalom, G. *J Am Acad Dermatol* 2017; **76**:662-9.e1. | Data not in extractable format |
| Strober, B. *J Am Acad Dermatol* 2017; **76**:432-40.e17. | Pooled data, all original studies already included |
| Strober, B. *J Am Acad Dermatol* 2017; **76**:655-61. | No outcomes not already extracted from Langley NEJM (2014) |
| Strober, B. *J Eur Acad Dermatol Venereol* 2018. | SR screened for additional papers – one identified Reich (2015) |
| Umezawa, Y. *J Eur Acad Dermatol Venereol* 2016; **30**:1957-60. | Inappropriate comparison: same biologic different treatment regimens |
| van de Kerkhof, P. *J Eur Acad Dermatol Venereol* 2017; **31**:477-82. | Outcome – serious infection not reported |
| Yiu, Z. Z. N. *Br J Dermatol* 2018. | Inappropriate study design – no comparator |
| Yiu, Z. Z. N. *J Invest Dermatol* 2018; **138**:534-41. | Data not in extractable format |

### Table S4. Risk of serious infections with biologic therapies at 10–16 weeks

| **Intervention** | **Peto OR (95% CI)** | **P value** | **$I^2$ value** |
|---|---|---|---|
| **Biologic vs. placebo** | | | |
| Etanercept | 0.56 (0.16 – 1.98) | 0.36 | 44% |
| Adalimumab | 1.24 (0.44 – 3.48) | 0.69 | 0% |
| Ustekinumab | 1.21 (0.44 – 3.35) | 0.71 | 13% |
| Secukinumab | 0.83 (0.20 – 3.56) | 0.81 | 0% |
| Ixekizumab | 1.77 (0.53 – 5.95) | 0.35 | 34% |
| Brodalumab | 1.80 (0.55 – 5.88) | 0.33 | 0% |
| Guselkumab | 1.00 (0.10 – 10.07) | 1.00 | 0% |
| Certolizumab pegol | 4.19 (0.56 – 31.11) | 0.16 | 0% |
| Tildrakizumab | 0.91 (0.10 – 8.74) | 0.94 | 0% |
| Risankizumab | 3.82 (0.40 – 36.80) | 0.25 | 0% |
| **Biologic vs. methotrexate in ≥4 to <18 years** | | | |
| Adalimumab | 4.40 (0.07 – 289.05) | 0.49 | NA |

| | | | |
|---|---|---|---|
| **Biologic vs. biologic** | | | |
| Brodalumab vs. ustekinumab | 1.33 (0.34 – 5.20) | 0.68 | NA |
| Risankizumab vs. ustekinumab | 0.26 (0.05 – 1.30) | 0.10 | 64% |
| Guselkumab vs. adalimumab | 0.32 (0.06 – 1.63) | 0.17 | 1% |
| Certolizumab pegol vs. etanercept | 4.51 (0.07 – 285.86) | 0.48 | NA |
| Tildrakizumab vs. etanercept | 4.50 (0.07 – 286.06) | 0.48 | NA |

**Abbreviations:** CI – confidence interval; NA – not applicable; OR - odds ratio

### Table S5. Risk of serious infections with biologic therapies at 1-year

| Comparison | Peto OR (95% CI) | P value |
|---|---|---|
| Guselkumab vs. adalimumab | 0.68 (0.12 – 3.93) | 0.66 |
| Risankizumab vs. ustekinumab | 0.06 (0.01 – 0.59) | 0.02 |

**Abbreviations:** CI – confidence interval; OR – odds ratio

**Table S6. Risk of bias for individual included studies**

| Study | Overall risk of bias | Risk of selection bias | Risk of performance bias | Risk of attrition bias | Risk of detection / measurement bias | Risk of outcome reporting bias | Risk of other bias |
|---|---|---|---|---|---|---|---|
| Asahina *et al.* 2010 | High | Low | High | Low | Low | Low | Low |
| Bachelez *et al.* 2015 | Low | Low | Low | Low | Low | Low | Low |
| Bagel *et al.* 2012 | Low | Low | Low | Low | Low | Low | Low |
| Blauvelt *et al.* 2015 | Low | Low | Low | Low | Low | Low | Low |
| Blauvelt *et al.* 2017 | Low | Low | Low | Low | Low | Low | Low |
| Cai *et al.* 2016 | Low | Low | Low | Low | Low | Low | Low |
| Gordon *et al.* 2006 | Very high | High | High | Low | High | Low | Low |
| Gordon *et al.* 2015 | Very high | Low | Very high | Low | Low | Low | Low |
| Gordon *et al.* 2016 | High | Low | High | Low | Low | Low | Low |
| Gordon *et al.* 2018 | Low | Low | Low | Low | Low | Low | Low |
| Gottlieb *et al.* 2011 | Very high | High | High | Low | Low | Low | Low |
| Gottlieb *et al.* 2018 | Low | Low | Low | Low | Low | Low | Low |
| Griffiths *et al.* 2010 | Low | Low | Low | Low | Low | Low | Low |
| Griffiths *et al.* 2015 | Low | Low | Low | Low | Low | Low | Low |
| Igarashi *et al.* 2011 | Very high | High | Low | High | Very high | Low | Low |
| Landells *et al.* 2015 | High | Low | Low | Low | Low | Low | Low |
| Langley *et al.* 2014 | Low | Low | Low | Low | Low | Low | Low |
| Lebwohl *et al.* 2015 | Low | Low | Low | Low | Low | Low | Low |
| Lebwohl *et al.* 2018 | Low | Low | Low | Low | Low | Low | Low |
| Leonardi *et al.* 2008 | Low | Low | Low | Low | Low | Low | Low |
| Leonardi *et al.* 2012 | High | Low | High | Low | Low | Low | Low |
| Menter *et al.* 2008 | Very high | Low | High | High | High | Low | Low |
| Paller *et al.* 2008 | Low | Low | Low | Low | Low | Low | Low |
| Papp *et al.* 2005 | Low | Low | Low | Low | Low | Low | Low |
| Papp *et al.* 2008 | Low | Low | Low | Low | Low | Low | Low |
| Papp *et al.* 2015 | Low | Low | Low | Low | Low | Low | Low |
| Papp *et al.* 2016 | Low | Low | Low | Low | Low | Low | Low |
| Papp *et al.* 2017 | High | Low | High | Low | Low | low | Low |
| Papp *et al.* Lancet 2017 | Low | Low | Low | Low | Low | Low | Low |
| Paul *et al.* 2015 | Low | Low | Low | Low | Low | Low | Low |
| Reich *et al.* 2012 | Low | Low | Low | Low | Low | Low | Low |
| Reich *et al.* JAAD 2017 | Low | Low | Low | Low | Low | Low | Low |
| Reich *et al.* JEADV 2017 | Low | Low | Low | Low | Low | Low | Low |
| Reich *et al.* Lancet 2017 | Low | Low | Low | Low | Low | Low | Low |
| Rich *et al.* 2013 | Low | Low | Low | Low | Low | Low | Low |
| Saurat *et al.* 2008 | Low | Low | Low | Low | Low | Low | Low |
| Strober *et al.* 2011 | Very high | High | High | Low | Low | Low | Low |
| Thaci *et al.* 2015 | High | Low | Low | Low | Low | Low | Low |

| Study | Overall risk of bias | Risk of selection bias | Risk of performance bias | Risk of attrition bias | Risk of detection / measurement bias | Risk of outcome reporting bias | Risk of other bias |
|---|---|---|---|---|---|---|---|
| Tsai *et al.* 2011 | High | Low | Low | Low | Low | Low | Low |
| Tyring *et al.* 2006 | Low | Low | Low | Low | Low | Low | Low |
| Van de Kerkhof *et al.* 2008 | Low | Low | Low | Low | Low | Low | Low |
| Yang *et al.* 2012 | High | High | Low | Low | Low | Low | Low |
| Zhu *et al.* 2013 | Very high | High | Low | Low | High | Low | Low |

N.B. High risk of bias is defined by either high risk of bias for one domain or an almost high risk of bias (not shown) for two domains

### Table S7. GRADE assessment profile table for Biologic vs. Placebo – Serious infection at week 10–16

| Quality assessment | | | | | | | No of patients | | Effect | | Certainty | Importance |
|---|---|---|---|---|---|---|---|---|---|---|---|---|
| **No of studies** | **Design** | **Risk of bias** | **Inconsistency** | **Indirectness** | **Imprecision** | **Other considerations** | **Biologic** | **Placebo - Serious infection at week 10-16 in adults** | **Relative (95% CI)** | **Absolute** | | |
| **Biologic vs Placebo - Etanercept** | | | | | | | | | | | | |
| 12 | randomised trials | no serious risk of bias | no serious inconsistency | no serious indirectness | very serious[1] | none | 6/2779 (0.2%) | 5/1514 (0.3%) | OR 0.56 (0.16 to 1.98) | 1 fewer per 1000 (from 3 fewer to 3 more) | ⊕⊕OO LOW | IMPORTANT |
| | | | | | | | | 0% | | - | | |
| **Biologic vs Placebo - Infliximab** | | | | | | | | | | | | |
| 1 | randomised trials | serious[2] | [3] | no serious indirectness | [3] | none | 0/84 (0%) | 0/45 (0%) | not pooled | not pooled | not applicable | IMPORTANT |
| | | | | | | | | 0% | | not pooled | | |
| **Biologic vs Placebo - Adalimumab** | | | | | | | | | | | | |
| 8 | randomised trials | very serious[2] | no serious inconsistency | no serious indirectness | very serious[1] | none | 11/2103 (0.5%) | 5/1100 (0.5%) | OR 1.24 (0.44 to 3.48) | 1 more per 1000 (from 3 fewer to 11 more) | ⊕OOO VERY LOW | IMPORTANT |
| | | | | | | | | 0% | | - | | |
| **Biologic vs Placebo - Ustekinumab** | | | | | | | | | | | | |
| 9 | randomised trials | serious[2] | no serious inconsistency | no serious indirectness | very serious[1] | none | 10/2490 (0.4%) | 6/1743 (0.3%) | OR 1.21 (0.44 to 3.35) | 1 more per 1000 (from 2 fewer to 8 more) | ⊕OOO VERY LOW | IMPORTANT |
| | | | | | | | | 0.3% | | 1 more per 1000 (from 2 fewer to 7 more) | | |

| | | | | | | | | | | | | |
|---|---|---|---|---|---|---|---|---|---|---|---|---|
| **Biologic vs Placebo - Secukinumab** | | | | | | | | | | | | |
| 5 | randomised trials | no serious risk of bias | no serious inconsistency | no serious indirectness | very serious[1] | none | 6/1720 (0.3%) | 3/761 (0.4%) | OR 0.83 (0.2 to 3.56) | 1 fewer per 1000 (from 3 fewer to 10 more) | ⊕⊕OO LOW | IMPORTANT |
| | | | | | | | | 0.3% | | 1 fewer per 1000 (from 2 fewer to 8 more) | | |
| **Biologic vs Placebo - Ixekizumab** | | | | | | | | | | | | |
| 4 | randomised trials | no serious risk of bias | no serious inconsistency | no serious indirectness | very serious[1] | none | 13/2449 (0.5%) | 3/819 (0.4%) | OR 1.28 (0.4 to 4.17) | 1 more per 1000 (from 2 fewer to 11 more) | ⊕⊕OO LOW | IMPORTANT |
| | | | | | | | | 0.3% | | 1 more per 1000 (from 2 fewer to 9 more) | | |
| **Biologic vs Placebo - Brodalumab** | | | | | | | | | | | | |
| 3 | randomised trials | no serious risk of bias | no serious inconsistency | no serious indirectness | very serious[1] | none | 14/2916 (0.5%) | 2/844 (0.2%) | OR 1.8 (0.55 to 5.88) | 2 more per 1000 (from 1 fewer to 11 more) | ⊕⊕OO LOW | IMPORTANT |
| | | | | | | | | 0.3% | | 2 more per 1000 (from 1 fewer to 14 more) | | |
| **Biologic vs Placebo - Guselkumab** | | | | | | | | | | | | |
| 3 | randomised trials | no serious risk of bias | no serious inconsistency | no serious indirectness | very serious[1] | none | 3/1033 (0.3%) | 1/464 (0.2%) | OR 1 (0.1 to 10.07) | 0 fewer per 1000 (from 2 fewer to 19 more) | ⊕⊕OO LOW | IMPORTANT |
| | | | | | | | | 0% | | - | | |
| **Biologic vs Placebo - Certolizumab pegol** | | | | | | | | | | | | |
| 4 | randomised trials | no serious risk of bias | no serious inconsistency | no serious indirectness | very serious[1] | none | 5/810 (0.6%) | 0/216 (0%) | OR 4.19 (0.56 to 31.11) | - | ⊕⊕OO LOW | IMPORTANT |
| | | | | | | | | 0% | | - | | |
| **Biologic vs Placebo - Tildrakizumab** | | | | | | | | | | | | |
| 3 | randomised trials | no serious risk of bias | no serious inconsistency | no serious indirectness | no serious imprecision | none | 4/1547 (0.3%) | 1/357 (0.3%) | OR 0.91 (0.1 to 8.74) | 0 fewer per 1000 (from 3 fewer to 21 more) | ⊕⊕⊕⊕ HIGH | IMPORTANT |
| | | | | | | | | 0% | | - | | |
| **Biologic vs Placebo - Risankizumab** | | | | | | | | | | | | |
| 2 | randomised trials | no serious risk of bias | no serious inconsistency | no serious indirectness | very serious[1] | none | 4/598 (0.7%) | 0/200 (0%) | OR 3.82 (0.4 to 36.8) | - | ⊕⊕OO LOW | IMPORTANT |
| | | | | | | | | 0% | | - | | |

[1] Downgraded by 1 increment if the confidence interval crossed one MID or by 2 increments if the confidence interval crossed both MIDs
[2] Downgraded by 1 increment if the majority of the evidence was at high risk of bias, and downgraded by 2 increments if the majority of the evidence was at very high risk of bias
[3] Unable to assess inconsistency, imprecision, or outcome due to lack of events in either arm

### Table S8. GRADE assessment profile table for Biologic vs. Placebo – Serious infection at week 10–16 in <18-year-olds

| Quality assessment | | | | | | | No of patients | | Effect | | Certainty | Importance |
|---|---|---|---|---|---|---|---|---|---|---|---|---|
| No of studies | Design | Risk of bias | Inconsistency | Indirectness | Imprecision | Other considerations | Biologic | Placebo - Serious infection at week 10–16 in <18-year-olds | Relative (95% CI) | Absolute | | |
| **Biologic vs Placebo** | | | | | | | | | | | | |
| 2 | randomised trials | no serious risk of bias | [1] | no serious indirectness | [1] | none | 0/179 (0%) | 0/142 (0%) | not pooled | not pooled | not applicable | IMPORTANT |
| | | | | | | | | 0% | | not pooled | | |
| **Biologic vs Placebo - Etanercept vs Placebo** | | | | | | | | | | | | |
| 1 | randomised trials | no serious risk of bias | [1] | no serious indirectness | [1] | none | 0/106 (0%) | 0/105 (0%) | not pooled | not pooled | not applicable | IMPORTANT |
| | | | | | | | | 0% | | not pooled | | |
| **Biologic vs Placebo - Ustekinumab vs Placebo** | | | | | | | | | | | | |
| 1 | randomised trials | no serious risk of bias | [1] | no serious indirectness | [1] | none | 0/73 (0%) | 0/37 (0%) | not pooled | not pooled | not applicable | IMPORTANT |
| | | | | | | | | 0% | | not pooled | | |

[1] Unable to assess inconsistency, imprecision, or outcome due to lack of events in either arm

### Table S9. GRADE assessment profile table for Biologic vs. Biologic – Serious infection at week 10–16

| Quality assessment | | | | | | | No of patients | | Effect | | Certainty | Importance |
|---|---|---|---|---|---|---|---|---|---|---|---|---|
| No of studies | Design | Risk of bias | Inconsistency | Indirectness | Imprecision | Other considerations | Biologic | Biologic - Serious infection at week 10-16 in adults | Relative (95% CI) | Absolute | | |
| **Ustekinumab vs Etanercept** | | | | | | | | | | | | |
| 1 | randomised trials | no serious risk of bias | not applicable | no serious indirectness | very serious[1] | none | 4/556 (0.7%) | 1/347 (0.3%) | OR 2.19 (0.36 to 13.31) | 3 more per 1000 (from 2 fewer to 34 more) | ⊕⊕OO LOW | IMPORTANT |
| | | | | | | | | 0.3% | | 4 more per 1000 (from 2 fewer to 36 more) | | |
| **Secukinumab vs Etanercept** | | | | | | | | | | | | |
| 1 | randomised trials | no serious risk of bias | not applicable | no serious indirectness | very serious[1] | none | 1/654 (0.2%) | 0/326 (0%) | OR 4.47 (0.07 to 286.66) | - | ⊕⊕OO LOW | IMPORTANT |
| | | | | | | | | 0% | | - | | |
| **Secukinumab vs Ustekinumab** | | | | | | | | | | | | |
| 1 | randomised trials | serious[2] | not applicable | no serious indirectness | very serious[1] | none | 1/337 (0.3%) | 2/339 (0.6%) | OR 0.52 (0.05 to 4.97) | 3 fewer per 1000 (from 6 fewer to 23 more) | ⊕OOO VERY LOW | IMPORTANT |
| | | | | | | | | 0.6% | | 3 fewer per 1000 (from 6 fewer to 23 more) | | |
| **Brodalumab vs. Ustekinumab** | | | | | | | | | | | | |
| 2 | randomised trials | no serious risk of bias | no serious inconsistency | no serious indirectness | very serious[1] | none | 11/2475 (0.4%) | 2/613 (0.3%) | OR 1.33 (0.34 to 5.2) | 1 more per 1000 (from 2 fewer to 13 more) | ⊕⊕OO LOW | IMPORTANT |
| | | | | | | | | 0.3% | | 1 more per 1000 (from 2 fewer to 12 more) | | |
| **Guselkumab vs. Adalimumab** | | | | | | | | | | | | |
| 3 | randomised trials | serious[2] | no serious inconsistency | no serious indirectness | very serious[1] | none | 3/1030 (0.3%) | 4/624 (0.6%) | OR 0.32 (0.06 to 1.63) | 4 fewer per 1000 (from 6 fewer to 4 more) | ⊕OOO VERY LOW | IMPORTANT |
| | | | | | | | | 0.6% | | 4 fewer per 1000 (from 6 fewer to 4 more) | | |
| **Certolizumab pegol vs Etanercept** | | | | | | | | | | | | |

| | | | | | | | | | | | | |
|---|---|---|---|---|---|---|---|---|---|---|---|---|
| 1 | randomised trials | no serious risk of bias | not applicable | no serious indirectness | very serious[1] | none | 1/332 (0.3%) | 0/168 (0%) | OR 4.51 (0.07 to 285.86) | - | ⊕⊕OO LOW | IMPORTANT |
| | | | | | | | | 0% | | - | | |
| **Tildrakizumab vs Etanercept** | | | | | | | | | | | | |
| 1 | randomised trials | no serious risk of bias | not applicable | no serious indirectness | very serious[1] | none | 1/621 (0.2%) | 0/313 (0%) | OR 4.5 (0.07 to 286.06) | - | ⊕⊕OO LOW | IMPORTANT |
| | | | | | | | | 0% | | - | | |
| **Risankizumab vs Ustekinumab** | | | | | | | | | | | | |
| 2 | randomised trials | no serious risk of bias | no serious inconsistency | no serious indirectness | no serious imprecision | none | 4/598 (0.7%) | 4/199 (2%) | OR 0.26 (0.05 to 1.3) | 15 fewer per 1000 (from 19 fewer to 6 more) | ⊕⊕⊕⊕ HIGH | IMPORTANT |
| | | | | | | | | 2% | | 15 fewer per 1000 (from 19 fewer to 6 more) | | |

[1] Downgraded by 1 increment if the confidence interval crossed one MID or by 2 increments if the confidence interval crossed both MIDs

[2] Downgraded by 1 increment if the majority of the evidence was at high risk of bias, and downgraded by 2 increments if the majority of the evidence was at very high risk of bias

### Table S10. GRADE assessment profile table for Biologic vs. Biologic – Serious infection at 1 year

| Quality assessment | | | | | | | No of patients | | Effect | | Certainty | Importance |
|---|---|---|---|---|---|---|---|---|---|---|---|---|
| No of studies | Design | Risk of bias | Inconsistency | Indirectness | Imprecision | Other considerations | Biologic | Biologic - Serious infection at 1 year in adults | Relative (95% CI) | Absolute | | |
| **Secukinumab vs Etanercept** | | | | | | | | | | | | |
| 1 | randomised trials | no serious risk of bias | not applicable | no serious indirectness | very serious[1] | none | 8/654 (1.2%) | 4/326 (1.2%) | OR 1 (0.3 to 3.34) | 0 fewer per 1000 (from 9 fewer to 28 more) | ⊕⊕OO LOW | IMPORTANT |
| | | | | | | | | 1.2% | | 0 fewer per 1000 (from 8 fewer to 27 more) | | |
| **Guselkumab vs. Adalimumab** | | | | | | | | | | | | |
| 1 | randomised trials | no serious risk of bias | not applicable | no serious indirectness | very serious[1] | none | 2/329 (0.6%) | 3/333 (0.9%) | OR 0.68 (0.12 to 3.93) | 3 fewer per 1000 (from 8 fewer to 25 more) | ⊕⊕OO LOW | IMPORTANT |
| | | | | | | | | 0.9% | | 3 fewer per 1000 (from 8 fewer to 25 more) | | |
| **Risankizumab vs Ustekinumab** | | | | | | | | | | | | |

| | | | | | | | | | | | | |
|---|---|---|---|---|---|---|---|---|---|---|---|---|
| 1 | randomised trials | serious[2] | not applicable | no serious indirectness | no serious imprecision | none | 1/126 (0.8%) | 3/40 (7.5%) | OR 0.06 (0.01 to 0.59) | 70 fewer per 1000 (from 29 fewer to 74 fewer) | ⊕⊕⊕O MODERATE | IMPORTANT |
| | | | | | | | | 7.5% | | 70 fewer per 1000 (from 29 fewer to 74 fewer) | | |

[1] Downgraded by 1 increment if the confidence interval crossed one MID or by 2 increments if the confidence interval crossed both MIDs

[2] Downgraded by 1 increment if the majority of the evidence was at high risk of bias, and downgraded by 2 increments if the majority of the evidence was at very high risk of bias

### Table S11. GRADE assessment profile table for Biologic vs. Methotrexate – Serious infection at 10–16 weeks in <18-year-olds

| Quality assessment | | | | | | | No of patients | | Effect | | Certainty | Importance |
|---|---|---|---|---|---|---|---|---|---|---|---|---|
| **No of studies** | **Design** | **Risk of bias** | **Inconsistency** | **Indirectness** | **Imprecision** | **Other considerations** | **Biologic** | **Methotrexate – Serious infections at week 10–16 in <18-year-olds** | **Relative (95% CI)** | **Absolute** | | |
| **Adalimumab vs Methotrexate** | | | | | | | | | | | | |
| 1 | randomised trials | no serious risk of bias | not applicable | no serious indirectness | very serious[1] | none | 1/77 (1.3%) | 0/37 (0%) | OR 4.4 (0.07 to 289.05) | - | ⊕⊕OO LOW | IMPORTANT |
| | | | | | | | | 0% | | - | | |

[1] Downgraded by 1 increment if the confidence interval crossed one MID or by 2 increments if the confidence interval crossed both MIDs

**Figure S1. Sensitivity analysis of serious infections for biologics vs. placebo at 10–16 weeks using Mantel-Haenszel methods (risk ratio) for fixed-effects models**

| Study or Subgroup | Biologic Events | Biologic Total | Placebo Events | Placebo Total | Weight | Risk Ratio M-H, Fixed, 95% CI |
|---|---|---|---|---|---|---|
| **1.1.1 Etanercept** | | | | | | |
| Bachelez Lancet 2015 | 2 | 336 | 0 | 108 | 6.9% | 1.62 [0.08, 33.43] |
| Bagel JAAD 2012 | 0 | 62 | 0 | 62 | | Not estimable |
| Gottlieb BJD 2011 | 1 | 141 | 0 | 68 | 6.2% | 1.46 [0.06, 35.32] |
| Griffiths Lancet 2015 UNCOVER-2 | 3 | 358 | 0 | 168 | 6.2% | 3.30 [0.17, 63.44] |
| Griffiths Lancet 2015 UNCOVER-3 | 0 | 382 | 2 | 193 | 30.3% | 0.10 [0.00, 2.10] |
| Lebwohl JAAD 2018 (200 & 400mg) | 0 | 168 | 0 | 57 | | Not estimable |
| Papp BJD 2005 | 0 | 390 | 1 | 193 | 18.3% | 0.17 [0.01, 4.04] |
| Reich JEADV 2017 (50mg/weekly) | 0 | 83 | 0 | 84 | | Not estimable |
| Reich Lancet 2017 reSURFACE2 | 0 | 313 | 1 | 156 | 18.3% | 0.17 [0.01, 4.07] |
| Strober BJD 2011 | 0 | 139 | 0 | 72 | | Not estimable |
| Tyring Lancet 2006 | 0 | 311 | 1 | 307 | 13.8% | 0.33 [0.01, 8.05] |
| Van de Kerkhof BJD 2008 | 0 | 96 | 0 | 46 | | Not estimable |
| **Subtotal (95% CI)** | | **2779** | | **1514** | **100.0%** | **0.54 [0.21, 1.42]** |
| Total events | 6 | | 5 | | | |
| Heterogeneity: Chi² = 4.62, df = 6 (P = 0.59); I² = 0% | | | | | | |
| Test for overall effect: Z = 1.25 (P = 0.21) | | | | | | |
| **1.1.2 Infliximab** | | | | | | |
| Yang CMJ 2012 | 0 | 84 | 0 | 45 | | Not estimable |
| **Subtotal (95% CI)** | | **84** | | **45** | | **Not estimable** |
| Total events | 0 | | 0 | | | |
| Heterogeneity: Not applicable | | | | | | |
| Test for overall effect: Not applicable | | | | | | |
| **1.1.3 Adalimumab** | | | | | | |
| Asahina JD 2010 | 1 | 123 | 0 | 46 | 8.6% | 1.14 [0.05, 27.42] |
| Blauvelt JAAD 2017 (100mg) | 2 | 333 | 0 | 174 | 7.8% | 2.62 [0.13, 54.27] |
| Cai JEADV 2016 | 0 | 338 | 0 | 87 | | Not estimable |
| Gordon JAAD 2006 | 1 | 96 | 0 | 52 | 7.7% | 1.64 [0.07, 39.54] |
| Gordon NEJM 2015 | 0 | 43 | 0 | 42 | | Not estimable |
| Menter JAAD 2008 | 5 | 814 | 4 | 398 | 64.0% | 0.61 [0.17, 2.26] |
| Reich JAAD 2017 (100mg) | 2 | 248 | 1 | 248 | 11.9% | 2.00 [0.18, 21.91] |
| Saurat BJD 2008 | 0 | 108 | 0 | 53 | | Not estimable |
| **Subtotal (95% CI)** | | **2103** | | **1100** | **100.0%** | **1.06 [0.42, 2.69]** |
| Total events | 11 | | 5 | | | |
| Heterogeneity: Chi² = 1.36, df = 4 (P = 0.85); I² = 0% | | | | | | |
| Test for overall effect: Z = 0.12 (P = 0.91) | | | | | | |
| **1.1.4 Ustekinumab** | | | | | | |
| Gordon Lancet 2018 (45mg & 150mg) UltIMMa-2 | 1 | 99 | 0 | 98 | 5.1% | 2.97 [0.12, 72.03] |
| Gordon Lancet 2018 (45mg & 150mg) UltIMMa-1 | 3 | 100 | 0 | 102 | 5.1% | 7.14 [0.37, 136.45] |
| Igarashi JD 2012 | 1 | 126 | 0 | 32 | 8.1% | 0.78 [0.03, 18.70] |
| Lebwohl NEJM 2015 AMAGINE-2 | 0 | 300 | 1 | 309 | 15.1% | 0.34 [0.01, 8.39] |
| Lebwohl NEJM 2015 AMAGINE-3 | 2 | 313 | 1 | 315 | 10.2% | 2.01 [0.18, 22.08] |
| Leonardi Lancet 2008 | 2 | 511 | 1 | 255 | 13.6% | 1.00 [0.09, 10.95] |
| Papp Lancet 2008 | 1 | 820 | 2 | 410 | 27.3% | 0.25 [0.02, 2.75] |
| Tsai JDS 2011 | 0 | 61 | 1 | 60 | 15.5% | 0.33 [0.01, 7.89] |
| Zhu JDD 2013 | 0 | 160 | 0 | 162 | | Not estimable |
| **Subtotal (95% CI)** | | **2490** | | **1743** | **100.0%** | **1.09 [0.47, 2.54]** |
| Total events | 10 | | 6 | | | |
| Heterogeneity: Chi² = 4.74, df = 7 (P = 0.69); I² = 0% | | | | | | |
| Test for overall effect: Z = 0.20 (P = 0.84) | | | | | | |
| **1.1.5 Secukinumab** | | | | | | |
| Blauvelt BJD 2015 | 2 | 118 | 1 | 59 | 21.7% | 1.00 [0.09, 10.81] |
| Langley NEJM 2014 ERASURE | 0 | 490 | 1 | 248 | 32.3% | 0.17 [0.01, 4.13] |
| Langley NEJM 2014 FIXTURE | 1 | 654 | 1 | 326 | 21.7% | 0.50 [0.03, 7.94] |
| Paul JEADV 2015 | 1 | 121 | 0 | 61 | 10.8% | 1.52 [0.06, 36.88] |
| Rich BJD 2012 | 2 | 337 | 0 | 67 | 13.5% | 1.01 [0.05, 20.72] |
| **Subtotal (95% CI)** | | **1720** | | **761** | **100.0%** | **0.68 [0.21, 2.25]** |
| Total events | 6 | | 3 | | | |
| Heterogeneity: Chi² = 1.19, df = 4 (P = 0.88); I² = 0% | | | | | | |
| Test for overall effect: Z = 0.63 (P = 0.53) | | | | | | |
| **1.1.6 Ixekizumab** | | | | | | |
| Gordon NEJM 2016 UNCOVER-1 | 4 | 865 | 0 | 431 | 14.3% | 4.49 [0.24, 83.20] |
| Griffiths Lancet 2015 UNCOVER-2 | 5 | 698 | 0 | 168 | 17.2% | 2.66 [0.15, 47.86] |
| Griffiths Lancet 2015 UNCOVER-3 | 4 | 771 | 2 | 193 | 68.5% | 0.50 [0.09, 2.71] |
| Leonardi NEJM 2012 | 0 | 115 | 0 | 27 | | Not estimable |
| **Subtotal (95% CI)** | | **2449** | | **819** | **100.0%** | **1.44 [0.43, 4.84]** |
| Total events | 13 | | 2 | | | |
| Heterogeneity: Chi² = 2.26, df = 2 (P = 0.32); I² = 11% | | | | | | |
| Test for overall effect: Z = 0.59 (P = 0.55) | | | | | | |
| **1.1.7 Brodalumab** | | | | | | |
| Lebwohl NEJM 2015 AMAGINE-2 | 4 | 1222 | 1 | 309 | 41.3% | 1.01 [0.11, 9.02] |
| Lebwohl NEJM 2015 AMAGINE-3 | 7 | 1253 | 1 | 315 | 41.4% | 1.76 [0.22, 14.25] |
| Papp BJD 2016 | 3 | 441 | 0 | 220 | 17.3% | 3.50 [0.18, 67.46] |
| **Subtotal (95% CI)** | | **2916** | | **844** | **100.0%** | **1.75 [0.47, 6.57]** |
| Total events | 14 | | 2 | | | |
| Heterogeneity: Chi² = 0.45, df = 2 (P = 0.80); I² = 0% | | | | | | |
| Test for overall effect: Z = 0.83 (P = 0.41) | | | | | | |
| **1.1.8 Guselkumab** | | | | | | |
| Blauvelt JAAD 2017 (100mg) | 0 | 329 | 0 | 174 | | Not estimable |
| Gordon NEJM 2015 | 2 | 208 | 0 | 42 | 38.3% | 1.03 [0.05, 21.05] |
| Reich JAAD 2017 (100mg) | 1 | 496 | 1 | 248 | 61.7% | 0.50 [0.03, 7.96] |
| **Subtotal (95% CI)** | | **1033** | | **464** | **100.0%** | **0.70 [0.09, 5.24]** |
| Total events | 3 | | 1 | | | |
| Heterogeneity: Chi² = 0.12, df = 1 (P = 0.73); I² = 0% | | | | | | |
| Test for overall effect: Z = 0.34 (P = 0.73) | | | | | | |
| **1.1.9 Certolizumab pegol** | | | | | | |
| Gottlieb JAAD 2018 (200 & 400mg) CIMPASI-2 | 1 | 178 | 0 | 49 | 34.0% | 0.84 [0.03, 20.26] |
| Gottlieb JAAD 2018 (200&400mg) CIMPASI-1 | 0 | 183 | 0 | 51 | | Not estimable |
| Lebwohl JAAD 2018 (200 & 400mg) | 1 | 332 | 0 | 57 | 37.1% | 0.52 [0.02, 12.67] |
| Reich BJD 2012 (200 & 400mg) | 3 | 117 | 0 | 59 | 28.9% | 3.56 [0.19, 67.79] |
| **Subtotal (95% CI)** | | **810** | | **216** | **100.0%** | **1.51 [0.28, 8.15]** |
| Total events | 5 | | 0 | | | |
| Heterogeneity: Chi² = 0.88, df = 2 (P = 0.64); I² = 0% | | | | | | |
| Test for overall effect: Z = 0.48 (P = 0.63) | | | | | | |
| **1.1.10 Tildrakizumab** | | | | | | |
| Papp BJD 2015 | 1 | 309 | 0 | 46 | 26.6% | 0.45 [0.02, 11.00] |
| Reich Lancet 2017 reSURFACE1 | 2 | 617 | 0 | 155 | 24.5% | 1.26 [0.06, 26.16] |
| Reich Lancet 2017 reSURFACE2 | 1 | 621 | 1 | 156 | 49.0% | 0.25 [0.02, 3.99] |
| **Subtotal (95% CI)** | | **1547** | | **357** | **100.0%** | **0.55 [0.11, 2.84]** |
| Total events | 4 | | 1 | | | |
| Heterogeneity: Chi² = 0.61, df = 2 (P = 0.74); I² = 0% | | | | | | |
| Test for overall effect: Z = 0.71 (P = 0.48) | | | | | | |
| **1.1.11 Risankizumab** | | | | | | |
| Gordon Lancet 2018 (45mg & 150mg) UltIMMa-2 | 3 | 294 | 0 | 98 | 50.0% | 2.35 [0.12, 45.08] |
| Gordon Lancet 2018 (45mg & 150mg) UltIMMa-1 | 1 | 304 | 0 | 102 | 50.0% | 1.01 [0.04, 24.68] |
| **Subtotal (95% CI)** | | **598** | | **200** | **100.0%** | **1.68 [0.20, 14.31]** |
| Total events | 4 | | 0 | | | |
| Heterogeneity: Chi² = 0.15, df = 1 (P = 0.70); I² = 0% | | | | | | |
| Test for overall effect: Z = 0.48 (P = 0.63) | | | | | | |

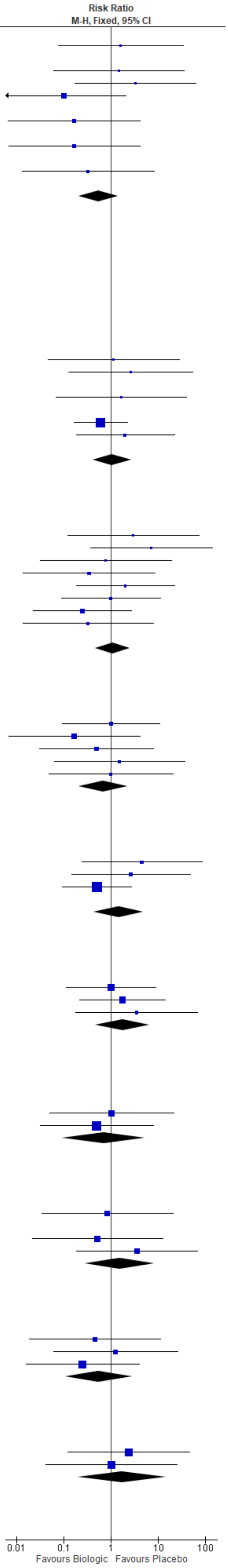

### Figure S2. Sensitivity analysis of serious infections for biologics vs. placebo at 10–16 weeks using Mantel-Haenszel methods (risk ratio) for random-effects models

| Study or Subgroup | Biologic Events | Biologic Total | Placebo Events | Placebo Total | Weight | Risk Ratio M-H, Random, 95% CI |
|---|---|---|---|---|---|---|
| **1.1.1 Etanercept** | | | | | | |
| Bachelez Lancet 2015 | 2 | 336 | 0 | 108 | 15.1% | 1.62 [0.08, 33.43] |
| Bagel JAAD 2012 | 0 | 62 | 0 | 62 | | Not estimable |
| Gottlieb BJD 2011 | 1 | 141 | 0 | 68 | 13.6% | 1.46 [0.06, 35.32] |
| Griffiths Lancet 2015 UNCOVER-2 | 3 | 358 | 0 | 168 | 15.8% | 3.30 [0.17, 63.44] |
| Griffiths Lancet 2015 UNCOVER-3 | 0 | 382 | 2 | 193 | 15.0% | 0.10 [0.00, 2.10] |
| Lebwohl JAAD 2018 (200 & 400mg) | 0 | 168 | 0 | 57 | | Not estimable |
| Papp BJD 2005 | 0 | 390 | 1 | 193 | 13.5% | 0.17 [0.01, 4.04] |
| Reich JEADV 2017 (50mg/weekly) | 0 | 83 | 0 | 84 | | Not estimable |
| Reich Lancet 2017 reSURFACE2 | 0 | 313 | 1 | 156 | 13.5% | 0.17 [0.01, 4.07] |
| Strober BJD 2011 | 0 | 139 | 0 | 72 | | Not estimable |
| Tyring Lancet 2006 | 0 | 311 | 1 | 307 | 13.5% | 0.33 [0.01, 8.05] |
| Van de Kerkhof BJD 2008 | 0 | 96 | 0 | 46 | | Not estimable |
| **Subtotal (95% CI)** | | **2779** | | **1514** | **100.0%** | **0.51 [0.16, 1.66]** |
| Total events | 6 | | 5 | | | |
| Heterogeneity: Tau² = 0.00; Chi² = 4.62, df = 6 (P = 0.59); I² = 0% | | | | | | |
| Test for overall effect: Z = 1.11 (P = 0.27) | | | | | | |
| **1.1.2 Infliximab** | | | | | | |
| Yang CMJ 2012 | 0 | 84 | 0 | 45 | | Not estimable |
| **Subtotal (95% CI)** | | **84** | | **45** | | **Not estimable** |
| Total events | 0 | | 0 | | | |
| Heterogeneity: Not applicable | | | | | | |
| Test for overall effect: Not applicable | | | | | | |
| **1.1.3 Adalimumab** | | | | | | |
| Asahina JD 2010 | 1 | 123 | 0 | 46 | 9.3% | 1.14 [0.05, 27.42] |
| Blauvelt JAAD 2017 (100mg) | 2 | 333 | 0 | 174 | 10.2% | 2.62 [0.13, 54.27] |
| Cai JEADV 2016 | 0 | 338 | 0 | 87 | | Not estimable |
| Gordon JAAD 2006 | 1 | 96 | 0 | 52 | 9.3% | 1.64 [0.07, 39.54] |
| Gordon NEJM 2015 | 0 | 43 | 0 | 42 | | Not estimable |
| Menter JAAD 2008 | 5 | 814 | 4 | 398 | 54.8% | 0.61 [0.17, 2.26] |
| Reich JAAD 2017 (100mg) | 2 | 248 | 1 | 248 | 16.4% | 2.00 [0.18, 21.91] |
| Saurat BJD 2008 | 0 | 108 | 0 | 53 | | Not estimable |
| **Subtotal (95% CI)** | | **2103** | | **1100** | **100.0%** | **1.00 [0.38, 2.64]** |
| Total events | 11 | | 5 | | | |
| Heterogeneity: Tau² = 0.00; Chi² = 1.36, df = 4 (P = 0.85); I² = 0% | | | | | | |
| Test for overall effect: Z = 0.00 (P = 1.00) | | | | | | |
| **1.1.4 Ustekinumab** | | | | | | |
| Gordon Lancet 2018 (45mg & 150mg) UltIMMa-2 | 1 | 99 | 0 | 98 | 9.5% | 2.97 [0.12, 72.03] |
| Gordon Lancet 2018 (45mg & 150mg) UltIMMa-1 | 3 | 100 | 0 | 102 | 11.1% | 7.14 [0.37, 136.45] |
| Igarashi JD 2012 | 1 | 126 | 0 | 32 | 9.6% | 0.78 [0.03, 18.70] |
| Lebwohl NEJM 2015 AMAGINE-2 | 0 | 300 | 1 | 309 | 9.5% | 0.34 [0.01, 8.39] |
| Lebwohl NEJM 2015 AMAGINE-3 | 2 | 313 | 1 | 315 | 16.9% | 2.01 [0.18, 22.08] |
| Leonardi Lancet 2008 | 2 | 511 | 1 | 255 | 16.9% | 1.00 [0.09, 10.95] |
| Papp Lancet 2008 | 1 | 820 | 2 | 410 | 16.9% | 0.25 [0.02, 2.75] |
| Tsai JDS 2011 | 0 | 61 | 1 | 60 | 9.6% | 0.33 [0.01, 7.89] |
| Zhu JDD 2013 | 0 | 160 | 0 | 162 | | Not estimable |
| **Subtotal (95% CI)** | | **2490** | | **1743** | **100.0%** | **0.97 [0.36, 2.61]** |
| Total events | 10 | | 6 | | | |
| Heterogeneity: Tau² = 0.00; Chi² = 4.74, df = 7 (P = 0.69); I² = 0% | | | | | | |
| Test for overall effect: Z = 0.05 (P = 0.96) | | | | | | |
| **1.1.5 Secukinumab** | | | | | | |
| Blauvelt BJD 2015 | 2 | 118 | 1 | 59 | 28.8% | 1.00 [0.09, 10.81] |
| Langley NEJM 2014 ERASURE | 0 | 490 | 1 | 248 | 16.0% | 0.17 [0.01, 4.13] |
| Langley NEJM 2014 FIXTURE | 1 | 654 | 1 | 326 | 21.3% | 0.50 [0.03, 7.94] |
| Paul JEADV 2015 | 1 | 121 | 0 | 61 | 16.1% | 1.52 [0.06, 36.88] |
| Rich BJD 2012 | 2 | 337 | 0 | 67 | 17.8% | 1.01 [0.05, 20.72] |
| **Subtotal (95% CI)** | | **1720** | | **761** | **100.0%** | **0.70 [0.19, 2.50]** |
| Total events | 6 | | 3 | | | |
| Heterogeneity: Tau² = 0.00; Chi² = 1.19, df = 4 (P = 0.88); I² = 0% | | | | | | |
| Test for overall effect: Z = 0.56 (P = 0.58) | | | | | | |
| **1.1.6 Ixekizumab** | | | | | | |
| Gordon NEJM 2016 UNCOVER-1 | 4 | 865 | 0 | 431 | 21.9% | 4.49 [0.24, 83.20] |
| Griffiths Lancet 2015 UNCOVER-2 | 5 | 698 | 0 | 168 | 22.3% | 2.66 [0.15, 47.86] |
| Griffiths Lancet 2015 UNCOVER-3 | 4 | 771 | 2 | 193 | 55.9% | 0.50 [0.09, 2.71] |
| Leonardi NEJM 2012 | 0 | 115 | 0 | 27 | | Not estimable |
| **Subtotal (95% CI)** | | **2449** | | **819** | **100.0%** | **1.17 [0.28, 4.88]** |
| Total events | 13 | | 2 | | | |
| Heterogeneity: Tau² = 0.20; Chi² = 2.26, df = 2 (P = 0.32); I² = 11% | | | | | | |
| Test for overall effect: Z = 0.22 (P = 0.83) | | | | | | |
| **1.1.7 Brodalumab** | | | | | | |
| Lebwohl NEJM 2015 AMAGINE-2 | 4 | 1222 | 1 | 309 | 37.9% | 1.01 [0.11, 9.02] |
| Lebwohl NEJM 2015 AMAGINE-3 | 7 | 1253 | 1 | 315 | 41.4% | 1.76 [0.22, 14.25] |
| Papp BJD 2016 | 3 | 441 | 0 | 220 | 20.7% | 3.50 [0.18, 67.46] |
| **Subtotal (95% CI)** | | **2916** | | **844** | **100.0%** | **1.65 [0.43, 6.32]** |
| Total events | 14 | | 2 | | | |
| Heterogeneity: Tau² = 0.00; Chi² = 0.45, df = 2 (P = 0.80); I² = 0% | | | | | | |
| Test for overall effect: Z = 0.72 (P = 0.47) | | | | | | |
| **1.1.8 Guselkumab** | | | | | | |
| Blauvelt JAAD 2017 (100mg) | 0 | 329 | 0 | 174 | | Not estimable |
| Gordon NEJM 2015 | 2 | 208 | 0 | 42 | 45.7% | 1.03 [0.05, 21.05] |
| Reich JAAD 2017 (100mg) | 1 | 496 | 1 | 248 | 54.3% | 0.50 [0.03, 7.96] |
| **Subtotal (95% CI)** | | **1033** | | **464** | **100.0%** | **0.70 [0.09, 5.35]** |
| Total events | 3 | | 1 | | | |
| Heterogeneity: Tau² = 0.00; Chi² = 0.12, df = 1 (P = 0.73); I² = 0% | | | | | | |
| Test for overall effect: Z = 0.35 (P = 0.73) | | | | | | |
| **1.1.9 Certolizumab pegol** | | | | | | |
| Gottlieb JAAD 2018 (200 & 400mg) CIMPASI-2 | 1 | 178 | 0 | 49 | 31.6% | 0.84 [0.03, 20.26] |
| Gottlieb JAAD 2018 (200&400mg) CIMPASI-1 | 0 | 183 | 0 | 51 | | Not estimable |
| Lebwohl JAAD 2018 (200 & 400mg) | 1 | 332 | 0 | 57 | 31.5% | 0.52 [0.02, 12.67] |
| Reich BJD 2012 (200 & 400mg) | 3 | 117 | 0 | 59 | 36.9% | 3.56 [0.19, 67.79] |
| **Subtotal (95% CI)** | | **810** | | **216** | **100.0%** | **1.23 [0.21, 7.37]** |
| Total events | 5 | | 0 | | | |
| Heterogeneity: Tau² = 0.00; Chi² = 0.88, df = 2 (P = 0.64); I² = 0% | | | | | | |
| Test for overall effect: Z = 0.23 (P = 0.82) | | | | | | |
| **1.1.10 Tildrakizumab** | | | | | | |
| Papp BJD 2015 | 1 | 309 | 0 | 46 | 29.1% | 0.45 [0.02, 11.00] |
| Reich Lancet 2017 reSURFACE1 | 2 | 617 | 0 | 155 | 32.2% | 1.26 [0.06, 26.16] |
| Reich Lancet 2017 reSURFACE2 | 1 | 621 | 1 | 156 | 38.7% | 0.25 [0.02, 3.99] |
| **Subtotal (95% CI)** | | **1547** | | **357** | **100.0%** | **0.50 [0.09, 2.80]** |
| Total events | 4 | | 1 | | | |
| Heterogeneity: Tau² = 0.00; Chi² = 0.61, df = 2 (P = 0.74); I² = 0% | | | | | | |
| Test for overall effect: Z = 0.78 (P = 0.43) | | | | | | |
| **1.1.11 Risankizumab** | | | | | | |
| Gordon Lancet 2018 (45mg & 150mg) UltIMMa-2 | 3 | 294 | 0 | 98 | 53.9% | 2.35 [0.12, 45.08] |
| Gordon Lancet 2018 (45mg & 150mg) UltIMMa-1 | 1 | 304 | 0 | 102 | 46.1% | 1.01 [0.04, 24.68] |
| **Subtotal (95% CI)** | | **598** | | **200** | **100.0%** | **1.59 [0.18, 13.94]** |
| Total events | 4 | | 0 | | | |
| Heterogeneity: Tau² = 0.00; Chi² = 0.15, df = 1 (P = 0.70); I² = 0% | | | | | | |
| Test for overall effect: Z = 0.42 (P = 0.67) | | | | | | |

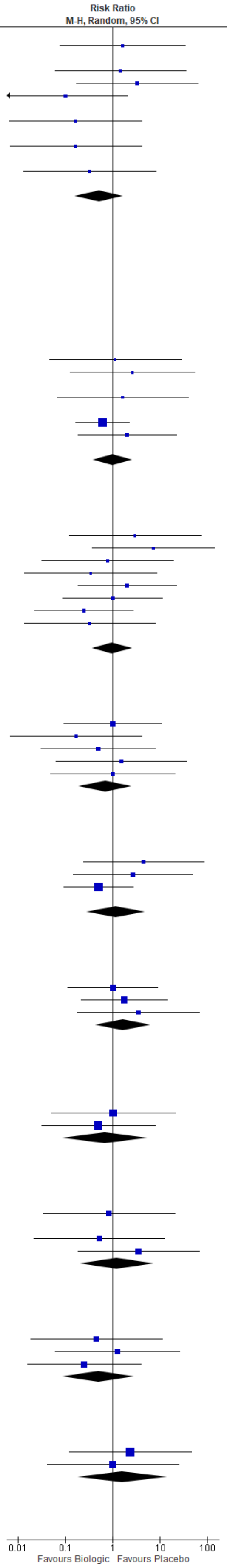
